# A newly identified pathology of Episodic Angioedema with Hypereosinophilia (Gleich’s Syndrome) revealed by Multi-Omics Analysis

**DOI:** 10.1101/2024.09.29.24313285

**Authors:** Tatsuya Koreeda, Hirokazu Muraoka, Yasunori Sato

## Abstract

Episodic Angioedema with Eosinophilia (Gleich syndrome) is a rare disease characterized by periodic angioedema, fever, and marked eosinophilia. This study aimed to elucidate the pathogenic factors of severe Gleich syndrome through comprehensive multi-omics analysis, including whole genome sequencing (WGS) and RNA sequencing (RNA-seq). A young female patient (age: 16-20 years old) presenting with periodic high fever, extensive urticaria/eczema, and marked eosinophilia was diagnosed with Gleich syndrome. The symptoms were severe, showing no spontaneous remission, but responded to oral steroid therapy with mild resolution. WGS of the patient’s blood identified high-impact pathogenic mutations in 16 genes, including PRDM16. RNA-seq analysis revealed differentially expressed genes (DEGs) associated with immune response regulation and viral defense. Combined z-score analysis of WGS and RNA-seq data highlighted ACE as a key gene, with its expression significantly downregulated during disease progression and recovered with treatment. IFNG was also identified. The findings suggest that decreased ACE expression, driven by PRDM16 mutations and altered IFNG expression, likely contributed to increased bradykinin levels and activation of the arachidonic acid cascade, resulting in the severe inflammation and angioedema characteristic of Gleich syndrome. This study highlights the utility of integrating WGS and RNA-seq data to uncover the molecular basis of rare diseases and provides a foundation for developing therapeutic strategies for hypereosinophilic syndromes.

## Introduction

Eosinophilia is defined as a condition characterized by an absolute eosinophil count in the peripheral blood exceeding 500 cells/µL [1,2]. Hypereosinophilia (HE) is defined as: 1) an absolute eosinophil count in the peripheral blood exceeding 1500 cells/µL in two blood samples taken at least 4 weeks apart; and/or 2) evidence of hypereosinophilia in tissues, which includes one or more of the following criteria: a) eosinophils constituting more than 20% of the total nucleated cells in the bone marrow; b) an abnormally intense eosinophilic infiltration in tissues as assessed by a pathologist; or c) extensive deposition of eosinophil granule proteins identified by special staining techniques [3]. Hypereosinophilic syndrome (HES) is defined by the following criteria: 1) the criteria for HE are met, 2) there is organ damage caused by peripheral tissue hypereosinophilia, and 3) other conditions that could cause organ damage have been excluded [3]. Another syndrome associated with HE is angioedema with eosinophilia, also known as Gleich syndrome. This rare clinical entity was first described by Gerald Gleich in 1984 and is characterized by angioedema with marked eosinophilia, accompanied by fever, periodic weight gain, and urticaria [4]. Hypereosinophilic syndrome (HES) was first proposed as a disease concept by Hardy and Anderson in 1968, with diagnostic criteria later developed by Chusid et al. in 1975. Initially, it was considered a syndrome encompassing many conditions diagnosed primarily by exclusion. Since then, the causes of some conditions previously classified as HES have been clarified [5,6], but many remain unexplained. The same applies to hypereosinophilia (HE), including Gleich syndrome. The same is true for HE, including Gleich syndrome. The low prevalence of HE and HES, combined with the need to initiate treatment before confirming strict diagnostic criteria, has left many aspects of these diseases medically unexplored.

In recent years, genomic analysis technologies, including Whole Genome Sequencing (WGS), Whole Exome Sequencing (WES), and targeted sequencing, have gained attention as research tools for elucidating the causes and mechanisms of diseases [7]. By comprehensively analyzing the entire spectrum of genetic variation, this technology can identify etiological genes and associated factors that are challenging to detect using conventional methods. WGS is particularly effective in the study of rare diseases, where many pathologies are strongly influenced by genetic factors [8–10]. In HE and HES, somatic mutations in haematopoietic cells have been investigated using targeted sequencing, as well as the relationship between clonal proliferation of eosinophils and novel disease-related mutations, using WES [11,12]. However, knowledge about the genetic background and mechanisms underlying the pathogenesis of HE and HES remains limited, as their causes are still unknown, and few cases have been analyzed using WGS. The aim of this study was to identify pathological factors in patients diagnosed with HE/Gleich syndrome of unknown etiology. We investigated the pathological factors of idiopathic hypereosinophilia by analyzing WGS and RNA-seq data obtained from these patients.

## Materials and Methods

### Patient

Patient with disease onset from April 2022 and diagnosed with severe Gleich syndrome in July 2023 was evaluated. The following clinical and laboratory information was collected: gender, age of onset, imaging evaluation of trunk organs and musculature, pathology of enlarged lymph nodes, pathology of skin lesions, peripheral blood counts including absolute eosinophil count, general biochemical tests, immunological tests, genetic tests, allergy tests, and parasite tests. The study protocol was approved by the Ethics Committees of the Clinic for Tamachi in Tokyo (approval number: CFT20230727), Japan, and all participants provided written informed consent.

### Sample Collection

Whole blood samples (2 mL) were collected from one patient and three healthy subjects. DNA and RNA were extracted simultaneously from the patient’s samples, which were collected at three time points: before treatment initiation, 30 days after treatment initiation, and 90 days after treatment initiation. The collected whole blood samples were used for subsequent WGS and RNA-seq analyses. The analysis workflow is provided in the Supplementary Information 1 (Figure S1).

### WGS Processing and Sequencing

Nucleic acids were extracted using the NucleoSpin RNA Blood Kit and NucleoSpin RNA/DNA Buffer Set from MACHEREY-NAGEL, following the manufacturer’s recommended protocol. In this process, genomic DNA was eluted first, followed by the extraction of total RNA. The extracted RNA was purified using the NucleoSpin RNA Clean-up XS Kit. The quality of the nucleic acid samples was assessed using Agilent Technologies’ TapeStation or BioAnalyzer. This assessment included double-stranded DNA quantification, agarose gel electrophoresis, and purification steps. The final quality assessment was based on the results from these instruments. Sequence libraries were prepared using Illumina’s TruSeq DNA PCR-Free Library Prep Kit and IDT for Illumina - TruSeq DNA UD Indexes v2. DNA was fragmented into hundreds of base pairs using Covaris Acoustic Solubilisers, followed by end-repair and phosphorylation. After size selection using magnetic beads, sequence libraries were constructed through 3’ dA tailing and ligation of indexed adaptors. Library quality was tested using Illumina’s MiSeq or iSeq systems to assess library size and concentration.

Sequence analysis was performed on Illumina’s NovaSeq X Plus system using the NovaSeq X Series 10B Reagent Kit and accompanying control software. Cluster generation and sequencing were conducted with Real Time Analysis (RTA) v4.6.2 and bclfastq2 v2.20 software.

### RNA-seq Processing and Sequencing

Nucleic acids were extracted from the provided samples using MACHEREY-NAGEL’s NucleoSpin RNA Blood Kit, and electrophoretic quantification was performed using a TapeStation or BioAnalyzer (Agilent Technologies) to assess the quality and quantity of the nucleic acids. Globin-depleted RNA samples were used, and adaptor sequences were added to both ends of the first-strand cDNA using the SMART method. PCR amplification was then performed using primers recognizing the adaptor sequence, and the PCR products were purified with AMPure XP (Beckman Coulter). The double-stranded cDNA was fragmented by a transposon-based tagging reaction, followed by the addition of adaptor sequences. Next, PCR amplification was carried out using indexed primers with distinct tag sequences to create a sequencing library. The quality of the library was assessed using an electrophoresis system. Reagent details for library preparation are provided in the data sheet. The processed samples underwent 150 base pair paired-end sequencing analysis using the NovaSeq system, and the raw data were converted to FASTQ file format using the provided software.

### Mapping Genomic Variation and Adding Annotation Information

Genomic mutation analysis was conducted using the DRAGEN Bio-IT Platform (Illumina). First, sequencing reads were mapped to the reference genome (hg38 decoy). Duplicates were marked, and these marked reads were used to identify candidate mutant bases, including single nucleotide substitutions and short insertions/deletions, that differ from the reference sequence. For multiple samples, the detection results were merged. The detected mutant bases were then filtered, and SnpEff and vcftools were used to assess the impact of the mutations. This assessment included comparing the candidate mutant bases with gene structure, predicting the impact of missense mutations (using FATHMM scores from dbNSFP), evaluating the mutation’s frequency across populations in gnomAD and East Asian allele frequencies, and adding clinical significance annotations from ClinVar.

### Variant Filtering and Analysis Workflow

Variant analysis was performed using SnpSift (version 4.3t) for filtering variants. Filtering criteria were restricted to variants with a total depth (DP) of 25 or greater to eliminate low-quality variants. Pathogenic SNVs were then extracted, focusing on those with annotation impacts labeled as HIGH and MODERATE. The filtered variants were analyzed to extract SNVs, Indels, and Structural Variants (SVs). The tools used for this analysis included AnnotSV [15], wANNOVAR [16], and SNPnexus [17]. AnnotSV used VCF files filtered by total depth and extracted variants with a class of 3 or higher in the ACMG guideline column. wANNOVAR focused on exonic regions, while SNPnexus excluded SNPs labeled ’Benign’ and ’Likely Benign’ in the ClinVar column. Data tables were generated from each tool, and columns without values were manually excluded upon inspection. Snowflake was utilized to examine the variants directly from the VCF files: after creating the database and schema in Snowflake, the VCF files were uploaded to an internal stage using SnowSQL (Version 1.2.32). The data were then loaded as semi-structured data into tables, and SQL was used for filtering.

### Mapping and Counting of Read Sequences

In the RNA sequencing analysis, read sequences were mapped to the human reference genome GRCh38 using Illumina’s NovaSeq Control Software version 1.7.5. Following mapping, quality control of the sequence data was performed using Illumina’s Real Time Analysis (RTA) software version 3.4.4. The resulting BCL files were converted to FASTQ format using bcl2fastq2 version 2.20. For mapping, a gene definition file based on GENCODE version 39 was used, incorporating detailed gene structures and annotation information. Annotation information was then added to the mapping data, and expression levels were organized in a tabular format. This process was conducted using Illumina’s DRAGEN Bio-IT Platform version 3.7.5, with the software’s default parameters applied at each step, from mapping read sequences to gene expression analysis. The resulting count data were used for subsequent gene expression analyses.

### Analysis of Established Gene Expression Levels Using RNA-Seq Data

Gene expression levels were analyzed using RNA-seq count data, focusing on genes labeled as ‘protein_coding’ before extracting Differentially Expressed Genes (DEGs). DEseq2 (Version 1.40.2) was used to identify DEGs by comparing the patient samples with those of three healthy subjects. Pathway enrichment analysis and protein-protein interaction (PPI) network analysis were performed on the identified DEGs. The pathway enrichment analysis was based on data from the GO term and KEGG databases. GO enrichment analysis classified the DEGs into three functional groups: biological processes, cellular components, and molecular functions [13]. The PPI network analysis was conducted using the STRING database in Cytoscape, with a Confidence Score cutoff of >0.7 and a maximum of 30 additional interactors. To identify hub genes, PPIs with a Confidence Score cutoff of >0.95 were screened, and the centrality of each gene in the PPI network was calculated using the CentiScaPe 2.2 plugin. Heatmaps were generated using the heatmap (Version 1.0.12) package.

### Integrated Variant and Gene Expression Analysis with Z-Score Standardization

After excluding low-quality variants and extracting those labeled as HIGH and MODERATE for annotation impact, the variants were matched with RNA-seq data based on transcript ID. Variants were then filtered to include only those with adjusted p-values less than 0.05 and an absolute Fold Change greater than 2. The PPI network centrality score was quantified using Centiscape 2.2. Following standardization by z-score based on adjusted p-values, absolute Fold Change, and centrality score values, the z-score totals were calculated for each gene.

### Ingenuity Pathway Analysis of RNA-Seq Data for Upstream and Causal Regulator Identification

Ingenuity Pathway Analysis (IPA) Expression Analysis RNA-seq data files were imported into IPA, with the log2FoldChange and padj columns mapped as ‘Expr Log Ratio’ and ‘Expr p-value,’ respectively. The analysis was performed with a Log Ratio cut-off range of -2.32 to 2.32 and an Expr p-value cut-off of 0.05. Within the Expression Analysis, Upstream Analysis was conducted to identify Upstream Regulators and Causal Regulators associated with the ACE gene. For Upstream Regulators, the absolute value of the Activation Z-score for each upstream gene was mapped to Target Molecules. For Causal Networks, intracellular pathway diagrams were created by generating networks for IFNG genes and selecting SubCellular layouts. Disease pathway information for Angioedema and Eosinophilia was also visualized under ‘Disease and Functions’.

## Results

### Clinical Findings

A young female patient (age: 16-20 years old) experienced periodic episodes of fever and myalgia, with temperatures ranging from 38-40°C, accompanied by chills and shivering, occurring one or two times a month since April 2022.

Around October of the same year, she developed edema, urticaria, and a generalized skin rash that left pigmentation. Blood tests from this period revealed markedly increased eosinophilia. During febrile episodes, the patient exhibited transient neutrophil predominance and significantly elevated CRP levels. Thorough examinations were conducted at the Clinic Fore Tamachi and Gunma University Hospital. A repeated clinical examinations revealed a total white blood cell count of 19,200/µL, with an eosinophil count of 16.4% (3148/µL). Hemoglobin and platelet counts were normal limits.

Total IgE was elevated at 504 IU/mL, but specific IgE levels for common grass and tree pollens, mites, animal dander, molds, and foods were within normal limits or negative. The patient, having been exposed to formalin, underwent a skin test for formalin sensitivity, which was negative. Screening for parasitic antibodies was also negative.

Rheumatoid factor, anti-nuclear antibodies, anti-ARS antibodies, MPO-ANCA, PR3-ANCA, IgG, IgA, IgM, C3, C4, and CH50 were all within normal limits. A peripheral blood smear examination revealed no morphological abnormalities. Lymphocyte surface marker tests showed the following: CD7 at 56.9%, CD3 at 44.7%, CD2 at 59.3%, CD4(-)CD8(+) at 9.3%, CD4(-)CD8(-) at 48.8%, CD4(+)CD8(+) at 41.2%, CD4(+)CD8(+) at 0.7%, and a CD4/CD8 ratio of 4.19.

Various imaging studies, including CT and MRI, did not reveal any tumorous lesions. However, imaging showed enlarged axillary lymph nodes, splenomegaly, myositis centered around both shoulders, and bilateral pleural effusions. The sIL-2R level was elevated at 1668 U/mL, but the pathology of the enlarged lymph nodes indicated dermatopathic lymphadenopathy, without evidence of lymphoma or malignancy, including immunostaining results. Pathological examination of the skin tissue raised a suspicion of vasculitis, though this was not definitive.

Genetic testing results were negative for the FIP1L1-PDGFRα fusion gene, PDGFB split signal, FGFR1 split signal, and MEFV gene mutation, with no clonal gene rearrangement detected in the T-cell receptor beta chain Cβ1. A summary of the clinical and laboratory information is provided below, and full test results are available in the Supplementary file (Supplementary Information 2).

Based on these results and the exclusion of autoimmune and hematological disorders and secondary eosinophilia, the patient was diagnosed with Episodic Angioedema with Hypereosinophilia (Gleich’s Syndrome). Treatment was initiated in July 2023 with oral glucocorticoids (prednisolone 30 mg/day), with subsequent tapering scheduled to the current maintenance dose of 1 mg/day. The patient achieved clinical and biological remission, generally by 30 days after treatment initiation and almost complete remission by 90 days.

### WGS-Based Identification of Patient-Specific Genetic Variants and Associated Mutations

Patient-specific genetic variants were analyzed using WGS for diagnostic purposes. Three variant analysis tools— AnnotSV, SNPnexus, and wANNOVAR — were employed. The AnnotSV analysis was filtered to include only variants classified with an ACMG classes of 3 or higher (Table 2). As a result, 26 variants with an ACMG class of 3 were identified, with no variants classified as more severe. Further filtering using a LOEUF_bin score of 0 identified deletion mutations in the FOXO1, PRDM16, SPAST, ITSN1, and DLGAP2 genes. Mutations in FOXO1 gene have been previously associated with rhabdomyosarcoma, a type of cancer. Variants in the PRDM16 gene have been linked to various cancers. Mutations in the SPAST gene are known to cause neurological disorders, particularly hereditary spastic paraplegia. However, no disease associations were identified for mutations in ITSN1 and DLGAP2 genes. In the SNPnexus-based analysis, the genes were listed as pathogenic according to ClinVar annotations (Table 3).

**Table 1.** Characteristics of the studied patient.

| Clinical Parameter | Patient's Data | Normal Range |
| --- | --- | --- |
| Gender | Female | - |
| Age of onset (years) | 16-20 | - |
| Age at diagnosis (years) | 16-20 | - |
| Symptoms | Angioedema<br>Urticaria<br>Pruritus<br>Periodic fever<br>Skin rash that leaves hyperpigmentation<br>Muscle pain<br>Splenomegaly<br>Lymphadenopathy | - |
| Serum total IgM (mg/dL) | 136 | 46 ~ 260 |
| Serum total IgG (mg/dL) | 1384 | 870 ~ 1700 |
| Serum total IgA (mg/dL) | 106 | 110 ~ 410 |
| Serum total IgE (IU/mL) | 504 | ≤170 |
| Aberrant T-cell population | No | - |
| CRP (mg/dL) | 17.2 | ≤0.14 |
| sIL-2R (U/mL) | 1668 | 157 ~ 474 |
| ANA (Titer) | <1:40 | <1:40 |
| MPO-ANCA (U/mL) | <1.0 | <3.5 |
| PR3-ANCA (U/mL) | <1.0 | <3.5 |
| C3 (mg/dL) | 114 | 86 ~ 160 |
| C4 (mg/dL) | 22 | 17 ~ 45 |
| anti-ARS antibodies (Index) | <5.0 | <25 |
| EBV DNA | Not detected | - |
| Trunk/Maxillofacial CT | Splenomegaly<br>Enlarged axillary lymph nodes | - |
| Trunk/Maxillofacial MRI | Suspected myositis around both shoulders<br>Reactive swelling of cervical and submandibular lymph nodes<br>Small amount of pleural effusion | - |
| Left axillary lymph node biopsy | Findings suggestive of dermatopathic lymphadenopathy<br>No lymphoma or malignant findings | - |
| Skin biopsy | Possible vasculitis (unclear) | - |
| FIP1L1-PDGFRα fusion gene | Negative | - |
| PDGFB split signal | Negative | - |
| FGFR1 split signal | Negative | - |
| MEFV gene mutations | Negative | - |

**Table 2.** Analysis of SVs Utilizing AnnotSV.

| Gene | Chr | Range | Loc | MOI | GenCC_disease | OMIM_phenotype |
| --- | --- | --- | --- | --- | --- | --- |
| FOXO1 | 13 | 40661542-40661624 | CDS |  | - | Alveolar<br>Rhabdomyosarcoma |
| PRDM16 | 1 | 3299300-3299421 | CDS | AD | dilated cardiomyopathy,<br>familial isolated dilated cardiomyopathy,<br>left ventricular noncompaction,<br>left ventricular noncompaction 8 | Dilated Cardiomyopathy,<br>Left ventricular<br>noncompaction 8 |
| PRDM16 | 1 | 3325997-3326054 | CDS | AD | dilated cardiomyopathy, familial isolated<br>dilated cardiomyopathy, left ventricular<br>noncompaction,<br>left ventricular noncompaction 8 | Dilated Cardiomyopathy<br>Left ventricular<br>noncompaction 8 |
| SPAST | 2 | 32111248-32111315 | CDS | AD;AR | Charlevoix-Saguenay spastic ataxia,<br>hereditary spastic paraplegia 4 | Spastic paraplegia 4 |
| ITSN1 | 21 | 33876194-33876246 | CDS |  | - | - |
| DLGAP2 | 8 | 999891-999994 | UTR | AD | complex neurodevelopmental disorder | - |

**Table 3.**
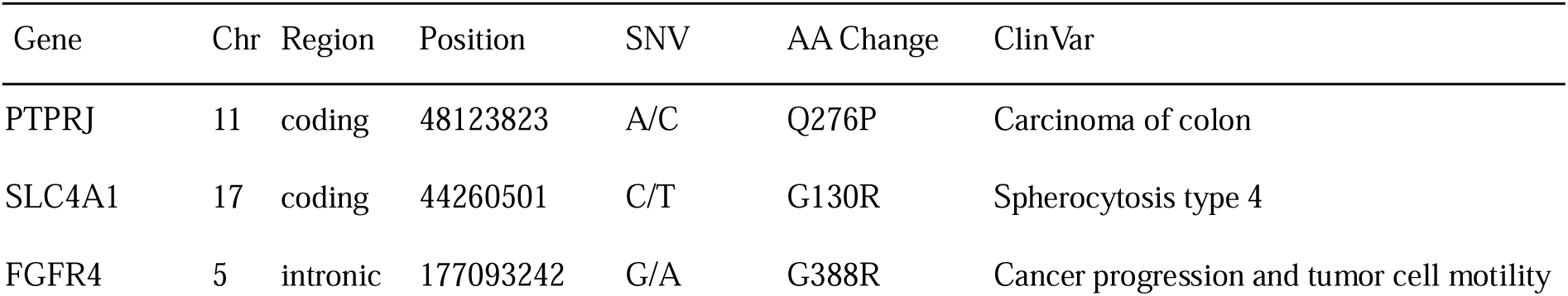
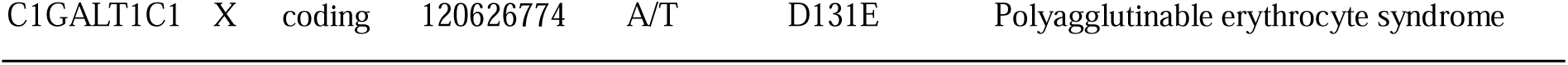
Analysis of SNVs Utilizing SNPnexus.

The analysis identified the following relevant mutations and corresponding genes: PTPRJ (rs1566734), SLC4A1 (rs121912749), FGFR4 (rs351855), and C1GALT1C1 (rs17261572). In PTPRJ, a non-synonymous SNV (A/C) was found at position 44,260,501 on chromosome 17, causing an amino acid change from Q276P and associated with carcinoma of the colon. In SLC4A1, a non-synonymous SNV (G/A) was observed in the intron region at position 177,093,242 on chromosome 5, resulting in an amino acid change to G388R, suggesting an association with cancer progression and tumor cell proliferation. A non-synonymous SNV (A/T) was identified in C1GALT1C1 at position 120,626,774 on the X chromosome, causing an amino acid change to D131E, and was associated with polyagglutinable erythrocyte syndrome.

In wANNOVAR, we identified a series of mutations across diverse genes, focusing on exonic regions (Table 4). As a result, we detected two distinct frameshift mutations across multiple exons (14, 16, and 17) in the APC gene. These mutations were identified as c.5400delT (p.N1800Kfs45), c.5454delT (p.N1818Kfs45), c.5399_5400del (p.N1800Kfs7), and c.5453_5454del (p.N1818Kfs7), all of which were heterozygous. In OR13C8, a c.242delT mutation in exon 1 (p.A83Pfs6) was observed in a homozygous form, and in PCDH17, a c.237_238del mutation (p.D80Qfs78) was found in exon 1, also in a homozygous form. For GSPT1, multiple frameshift mutations were identified in exon 15, including c.1902_1909del (p.K635Sfs63), c.1491_1498del (p.K498Sfs63), and c.1905_1912del (p.K636Sfs*63), all of which were homozygous. In MED1, two heterozygous frameshift mutations were identified in exon 17, c.3210delA (p.T1071Qfs2) and c.3210_3211del (p.T1071Sfs28). In SLC35G4, synonymous SNVs c.C570T and c.C570C were detected in exon 1, both heterozygous, indicating no change in the amino acid sequence of the protein (p.G190G). In exon 4 of MADCAM1, a non-synonymous SNV, c.T718C (p.S240P), was identified in a heterozygous form.

**Table 4.**
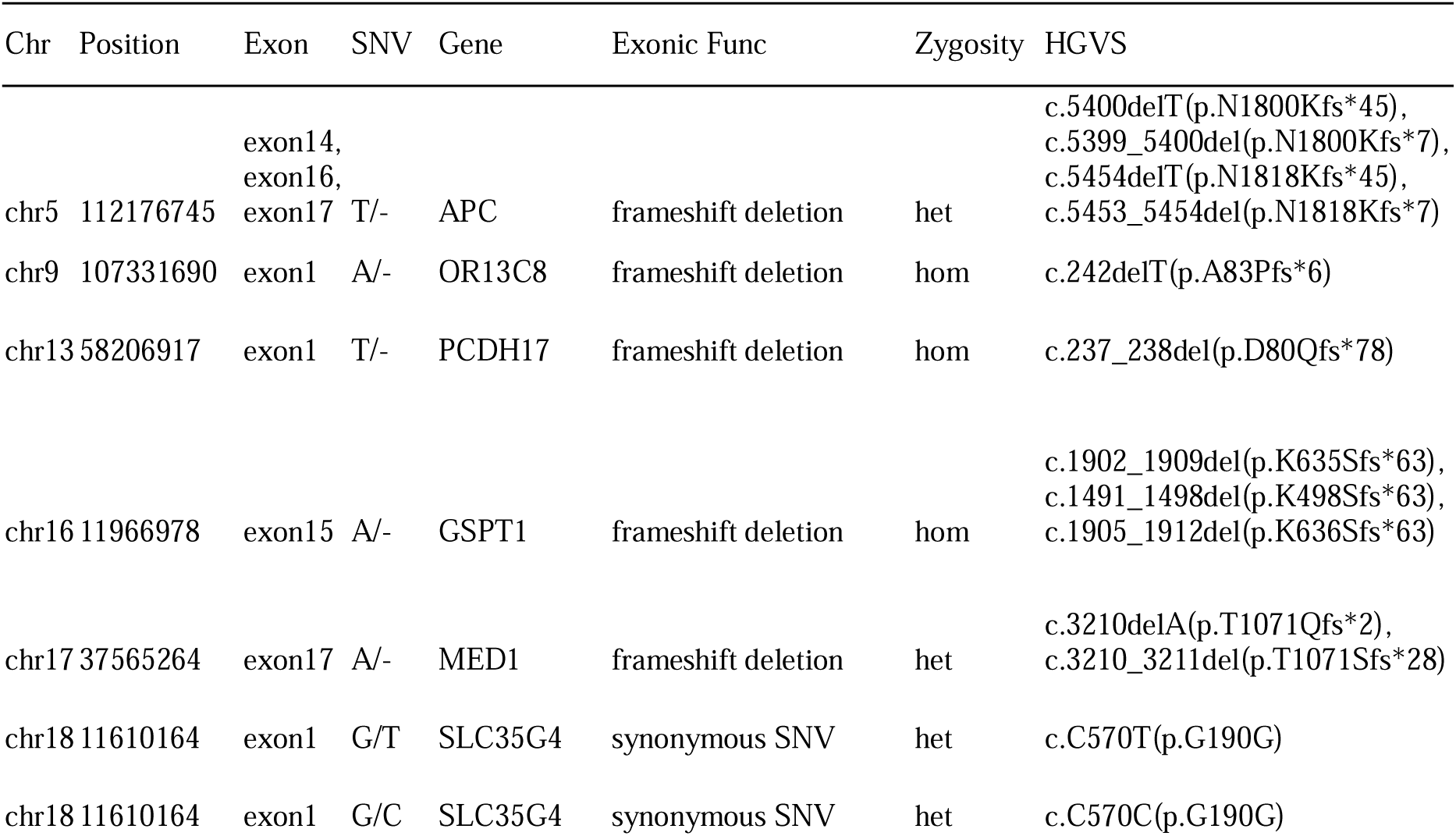
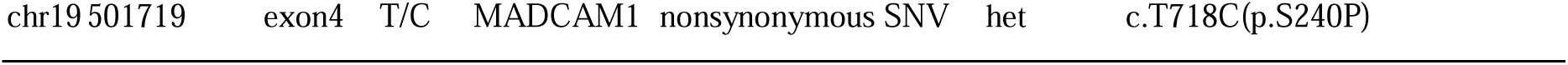
Analysis of SNVs Utilizing wANNOVAR.

### Enrichment Analysis and PPI Network Analysis Utilizing RNA-seq DEGs

The DEGs identified from the RNA-seq data in healthy subjects and patients were further analyzed using GO and KEGG pathway analyses. Upregulated genes were significantly enriched in the biological process categories of ‘regulation of response to biotic stimulus’ and ‘response to virus,’ as well as in the molecular function categories of ‘DNA-binding transcription factor binding’ and ‘carbohydrate binding.’ These genes were also significantly enriched in the cellular component categories of ‘endocytic vesicle’ and ‘specific granule’ (Figure 1a and Table 5). Conversely, downregulated genes were significantly enriched in the biological process categories of ‘leukocyte-mediated immunity’ and ‘cell killing,’ in the molecular function categories of ‘carbohydrate binding’ and ‘immune receptor activity,’ and in the cellular component categories of ‘external side of plasma membrane’ and ‘endoplasmic reticulum lumen’ (Figure 1b and Table 6). Additionally, KEGG pathway analysis revealed that the most significantly enriched gene pathways were those associated with the ‘PI3K-Akt signaling pathway’ and ‘Human papillomavirus infection’ (Figure 1c).

**Figure 1.**
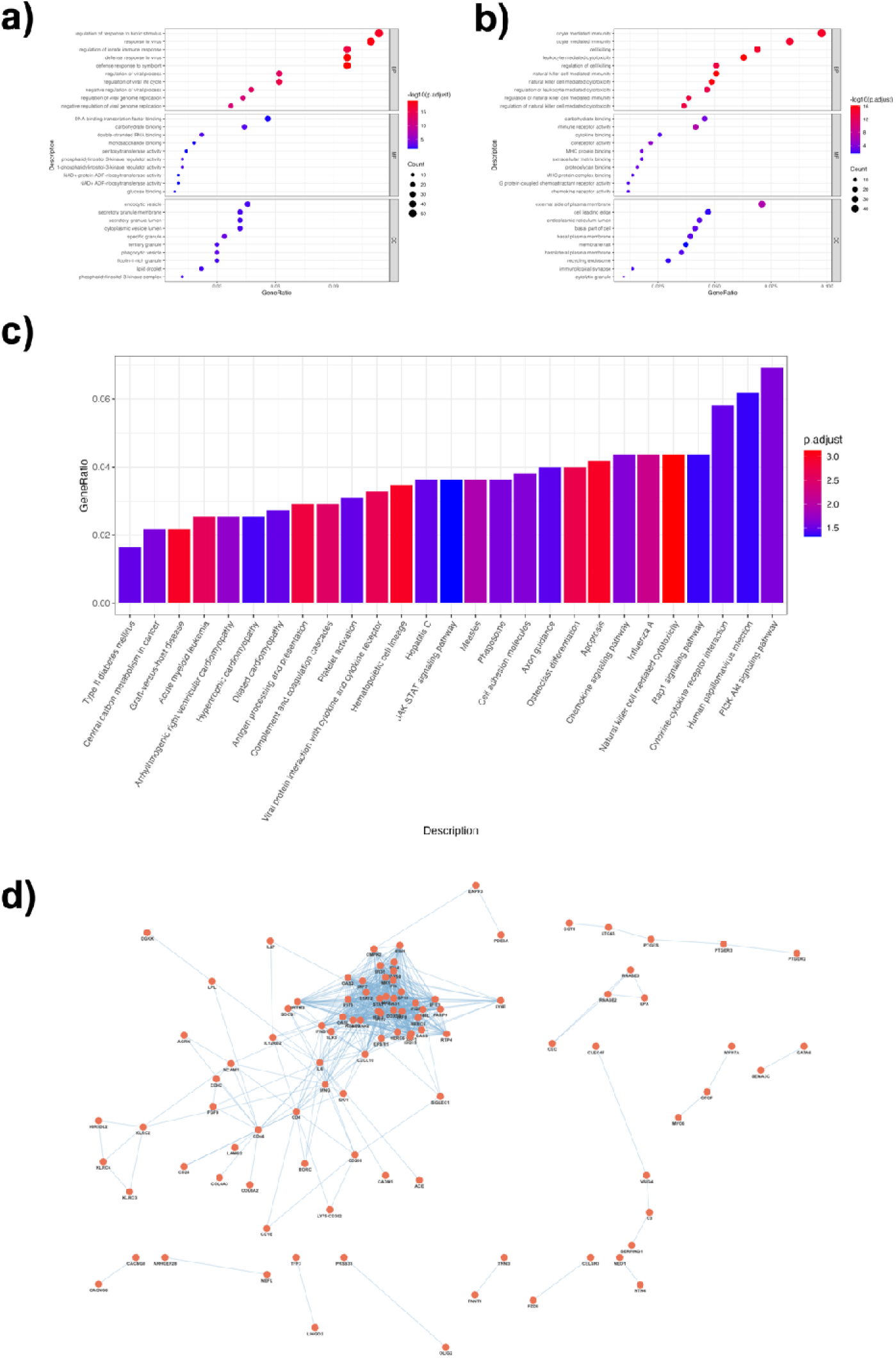
Pathway Enrichment Analysis Utilizing GO and KEGG, and PPI Network Analysis Pathway enrichment analysis using GO and KEGG terms, and PPI network analysis based on patient RNA-seq data. a) Pathway enrichment analysis using GO terms for DEGs of patient-derived upregulated genes in biological process (BP), molecular function (MF), and cellular component (CC) categories. The top 10 terms in each category are shown. b) Pathway enrichment analysis using GO terms for DEGs of patient-derived downregulated genes. c) Pathway enrichment analysis using KEGG terms for DEGs of patient-derived genes with variable expression. d) Pathway enrichment analysis using GO terms for DEGs of patient-derived genes with variable expression. The vertical axis represents the percentage of DEGs in each term. e) PPI network analysis using DEGs of patient-derived genes with variable expression. Red circles represent nodes, and blue lines represent edges.

**Table 5.** Enrichment Analysis Using Upregulated Genes Derived from RNA-seq Data.

| Category | Term | Description | Count | % | p.adjust |
| --- | --- | --- | --- | --- | --- |
| BP | GO:0051607 | defense response to virus | 47 | 9.69072165 | 8.65E-20 |
| BP | GO:0140546 | defense response to symbiont | 47 | 9.69072165 | 8.65E-20 |
| BP | GO:0009615 | response to virus | 53 | 10.9278351 | 3.64E-19 |
| BP | GO:0002831 | regulation of response to biotic stimulus | 55 | 11.3402062 | 3.14E-18 |
| BP | GO:0045088 | regulation of innate immune response | 47 | 9.69072165 | 2.09E-16 |
| BP | GO:1903900 | regulation of viral life cycle | 30 | 6.18556701 | 3.82E-16 |
| MF | GO:0003725 | double-stranded RNA binding | 11 | 2.2 | 0.00202485 |
| MF | GO:0030246 | carbohydrate binding | 22 | 4.4 | 0.00202485 |
| MF | GO:0046935 | 1-phosphatidylinositol-3-kinase regulator activity | 6 | 1.2 | 0.00202485 |
| MF | GO:0035014 | phosphatidylinositol 3-kinase regulator activity | 6 | 1.2 | 0.00389573 |
| MF | GO:0140297 | DNA-binding transcription factor binding | 28 | 5.6 | 0.01965112 |
| MF | GO:0005536 | glucose binding | 4 | 0.8 | 0.01965112 |
| CC | GO:0042581 | specific granule | 17 | 3.37301587 | 0.00036595 |
| CC | GO:0045335 | phagocytic vesicle | 15 | 2.97619048 | 0.00078833 |
| CC | GO:0070820 | tertiary granule | 15 | 2.97619048 | 0.00329658 |
| CC | GO:0030139 | endocytic vesicle | 23 | 4.56349206 | 0.00399692 |
| CC | GO:0005811 | lipid droplet | 11 | 2.18253968 | 0.00446821 |
| CC | GO:0005942 | phosphatidylinositol 3-kinase complex | 6 | 1.19047619 | 0.00446821 |

**Table 6.** Enrichment Analysis Using Downregulated Genes Derived from RNA-seq Data.

| Category | Term | Description | Count | % | p.adjust |
| --- | --- | --- | --- | --- | --- |
| BP | GO:0002228 | natural killer cell mediated immunity | 25 | 5.08130081 | 5.55E-17 |
| BP | GO:0001909 | leukocyte mediated cytotoxicity | 31 | 6.30081301 | 6.45E-17 |
| BP | GO:0042267 | natural killer cell mediated cytotoxicity | 24 | 4.87804878 | 1.16E-16 |
| BP | GO:0002443 | leukocyte mediated immunity | 48 | 9.75609756 | 1.62E-15 |
| BP | GO:0001906 | cell killing | 34 | 6.91056911 | 1.98E-15 |
| BP | GO:0002449 | lymphocyte mediated immunity | 41 | 8.33333333 | 3.27E-15 |
| MF | GO:0140375 | immune receptor activity | 21 | 4.1749503 | 1.84E-07 |
| MF | GO:0015026 | coreceptor activity | 11 | 2.18687873 | 1.79E-05 |
| MF | GO:0042287 | MHC protein binding | 9 | 1.78926441 | 0.00033639 |
| MF | GO:0030246 | carbohydrate binding | 23 | 4.57256461 | 0.00033639 |
| MF | GO:0043394 | proteoglycan binding | 8 | 1.59045726 | 0.00062812 |
| MF | GO:0050840 | extracellular matrix binding | 9 | 1.78926441 | 0.00189935 |
| CC | GO:0009897 | external side of plasma membrane | 36 | 7.10059172 | 2.48E-07 |
| CC | GO:0044194 | cytolytic granule | 5 | 0.98619329 | 0.00211125 |
| CC | GO:0045178 | basal part of cell | 21 | 4.14201183 | 0.00211125 |
| CC | GO:0009925 | basal plasma membrane | 20 | 3.94477318 | 0.00211125 |
| CC | GO:0005788 | endoplasmic reticulum lumen | 22 | 4.33925049 | 0.00211125 |
| CC | GO:0016323 | basolateral plasma membrane | 18 | 3.55029586 | 0.00310627 |

Next, a PPI network analysis was performed using the DEGs. In the PPI network, which consisted of 185 DEGs, 214 nodes and 676 edges were mapped based on protein-protein interactions with interaction scores greater than 0.7 (Figure 1d). Using an interaction score greater than 0.95 to identify hub genes, the most highly connected hub genes were RSAD2 (degree = 9), MX1 (degree = 8), ISG15 (degree = 8), and IFI44L (degree = 7). A heatmap of the DEGs and a volcano plot showing the time-series changes in each gene expression level are provided in the Supplementary Information 1 (Figure S2).

### Identification of Key Genes Utilizing Z-Score Analysis

Variants from WGS data labeled as HIGH in annotation impact, along with gene expression levels from RNA-seq data and centrality scores from PPI network analysis, were evaluated utilizing z-score analysis (Table 7). Among the Z-scores, IFI27 had the highest value (Z-Score: 4.47619923). OAS2 (Degree: 19) and ACE (ABS_log2FC: 9.10262032) exhibited the highest centrality score and absolute log2 fold change, respectively. The smallest adjusted p-value was observed for IFI27 (padj: 4.18E-65). The effects of the mutations were as follows: splice acceptor variant & intron variant in three cases, start lost & conservative inframe deletion in one case, frameshift variant in three cases, stop lost in one case, stop gained in two cases, and splice donor variant & intron variant in one case. Z-score values were utilized for GO term and KEGG pathway enrichment analysis as well as PPI network analysis (Figure S3). The plots of Effect Type count values for genomic variants in this patient are provided in the Supplementary Information (Figure S4).

**Table 7.** Identification of Key Disease-Associated Genes Based on Z-Score Values.

| Gene | POS | Variant | Effect | Degree | ABS_log2FC | padj | Z_Score |
| --- | --- | --- | --- | --- | --- | --- | --- |
| IFI27 | 14:94105764 | G/T | splice_acceptor_variant&intron_variant | 14 | 7.22991464 | 4.18E-65 | 4.47619973 |
| IFI27 | 14:94115784 | TGGCCAT<br>GGC/T | start_lost&conservative_inframe_deletion | 14 | 7.22991464 | 4.18E-65 | 4.47619973 |
| IFI44 | 1:78662885 | G/GT | frameshift_variant | 18 | 3.62858894 | 1.20E-30 | 3.63485564 |
| OAS2 | 12:113010483 | A/G | stop_lost | 19 | 2.51335138 | 7.25E-23 | 3.33028482 |
| EPSTI1 | 13:42888298 | T/TTCAGG | frameshift_variant | 17 | 3.22174861 | 6.81E-18 | 3.27200626 |
| ACE | 17:63486300 | C/A | stop_gained | 4 | 9.10262032 | 0.00107677 | 3.11768804 |
| MSR1 | 8:16186158 | C/T | splice_donor_variant&intron_variant | 4 | 3.28217749 | 5.35E-04 | 0.73409481 |
| COL6A2 | 21:46125455 | A/AC | splice_acceptor_variant&intron_variant | 1 | 3.28721175 | 7.06E-18 | 0.34946627 |
| SIGLEC10 | 19:51417359 | C/T | splice_acceptor_variant&intron_variant | 2 | 1.88542826 | 1.09E-08 | -0.0807563 |
| RETSAT | 2:85350974 | TA/T | frameshift_variant | 3 | 1.48768267 | 2.26E-04 | -0.1410527 |
| CC2D2A | 4:15480736 | C/T | stop_gained | 1 | 1.84782388 | 4.07E-04 | -0.4083765 |

### Dynamics of Gene Expression in Pre- and Post-Treatment Time Series

The temporal changes in expression levels for selected genes were assessed. For each gene group, log2 fold changes were calculated at three time points: pre-treatment, 1 month, and 3 months post-treatment. Significant changes in expression levels were observed when comparing pre-treatment samples to those from healthy subjects, particularly for the genes ACE, CC2D2A, COL6A2, EPST11, IFI27, IFI44, MSR1, OAS2, RETSAT, and SIGLEC10 (Figure 2a). Overall, many genes exhibited significant temporal changes in expression levels compared to pre-treatment. For example, EPST11 was strongly upregulated before treatment, with this upregulation alleviated after one month and further reduced after three months. A similar pattern was observed for IFI27, IFI44, and CC2D2A. Conversely, ACE and RETSAT exhibited a reverse trend, with marked suppression of expression before and one month post-treatment, followed by recovery after three months. PTPRJ and SIGLEC10 showed reduced expression at 1 and 3 months post-treatment compared to healthy subjects, while OR13C8 and SLC35G had a read count of 0 in both patients and healthy subjects. Enrichment analysis revealed that GO terms related to the regulation of immune response, leukocyte-mediated immunity, and cell killing were enriched in pre-treatment samples but were no longer enriched at 1 and 3 months post-treatment (Figure 2b). In contrast, GO terms involved in chemotaxis, cell migration, and cell differentiation were newly enriched at 1 and 3 months post-treatment. The pre-treatment samples were plotted in the third quadrant, while the 1- and 3-month post-treatment samples were plotted in the first quadrant (Figure 2c). A heatmap identified genes with high loading on the negative axis of PC1, including BFAR, XG, RCE1, and COL18A1 (Figure 2d). For each gene showing a recovery trend after 3 months, the OMIM database was used to search for causative genes with phenotypes similar to those observed in the patient. The results indicated that a decrease in ACE led to an increase in bradykinin, contributing to the development of angioedema [14,15].

**Figure 2.**
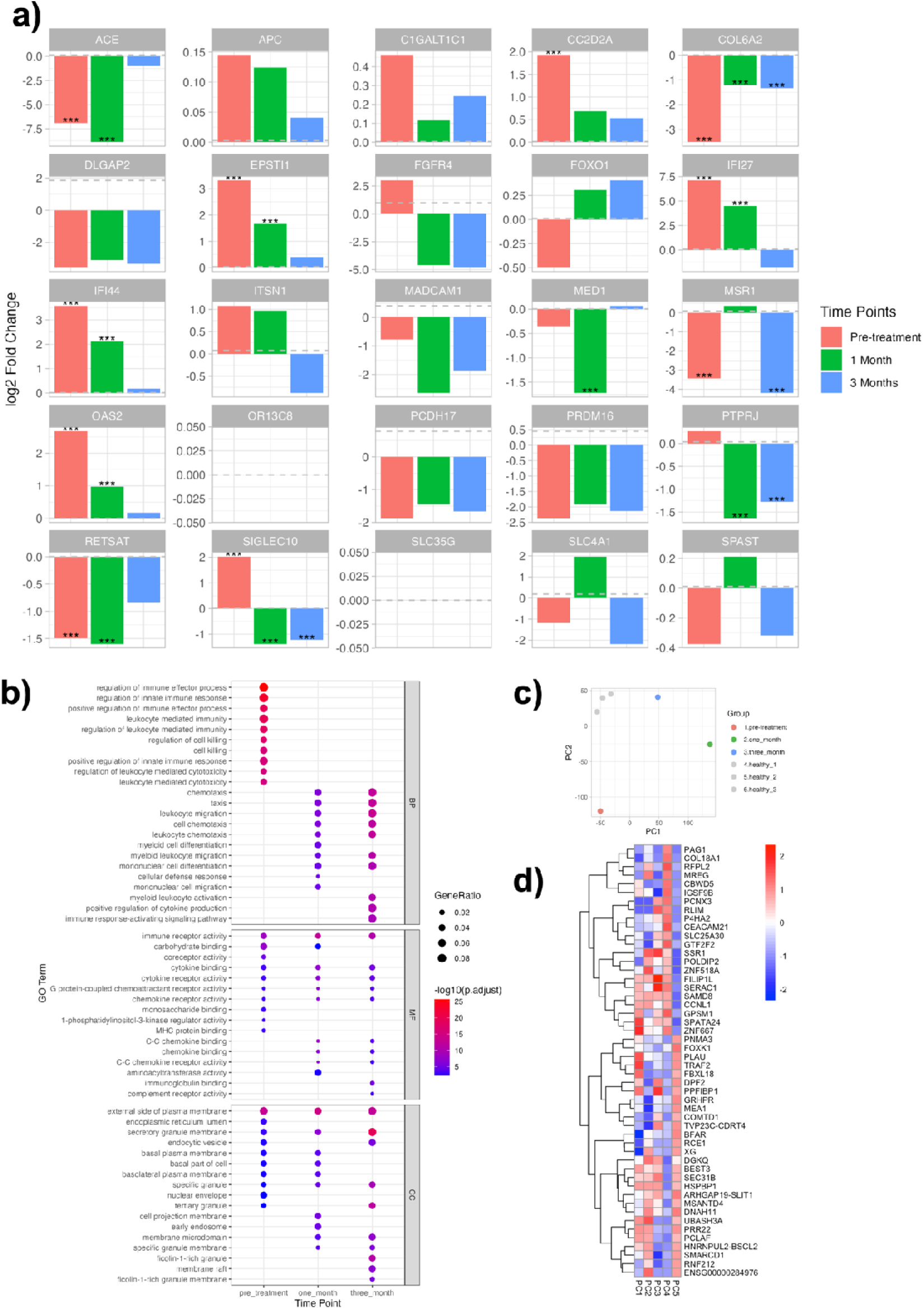
Analysis of Time-Series Changes in Gene Expression Levels and Pathway Enrichment Analysis of Changes in Gene Expression Levels and Enriched Pathways Using Patient Samples Collected at Pre-treatment, 1 Month, and 3 Month Time Points. a) Comparison of expression levels of candidate target genes at each time point. Three asterisks indicate an adjusted p-value of less than 0.01, representing a significant difference from the healthy group. b) Transition of enriched GO terms at each time point, representing Biological Process (BP), Molecular Function (MF), and Cellular Component (CC). c) PCA plot using RNA-seq data from healthy subjects and patients at each time point. d) PCA for each gene. e) Heatmap displaying loading values, showing PC1 to PC5, clustered in rows.

### Exploration of upstream and master regulators of the ACE gene by IPA

Analysis of the dynamics of gene expression in pre- and post-treatment time series suggested a relationship between the ACE gene and patients with the disease. To further explore this, we investigated the relationship between the ACE gene and the disease by visualizing upstream regulators and causal networks using IPA for the ACE gene. When we searched for upstream regulators of ACE using pre-treatment RNA-seq data, we found that the IFNG gene had the highest Activation z-score of 4.403 (Figure S5a). Similarly, in the causal network’s Activation z-scores, the IFNG gene also had the highest Activation z-score of 4.217 (Figure S5b). The network visualization showed an interaction from IFNG in the extracellular space to ACE in the plasma membrane (Figure S5c). To further investigate the association between IFNG and this patient, a drill-down analysis using IPA for the IFNG gene identified an association with idiopathic eosinophilia [16]. Pathways enriched in IPA for this disease, including pathway diagrams for RNA virus infection and network diagrams for Angioedema, which are suggested to be relevant to the disease pathogenesis, are provided in the Supplementary Information (Figures S6, 7).

## Discussion

The aim of this study was to identify the pathological factors in patients diagnosed with HE/Gleich syndrome of unknown etiology using WGS and RNA-seq analysis.

The patient presented with clinical symptoms consistent with Gleich’s syndrome, including marked eosinophilia exceeding 5000/µL at its peak, periodic fever, and recurrent edema and urticaria. However, the patient also exhibited a generalized skin rash with pathological findings of vasculitis and pigmentation, transient neutrophil-predominant leukemoid reactions associated with fever, markedly elevated CRP levels, and splenomegaly. These findings are not commonly observed in Gleich’s syndrome, but similar cases have been reported [17–19]. Although this patient did not meet the strict diagnostic criteria for HES, as eosinophilic infiltration was not clearly demonstrated in various organ assessments, some consider Gleich syndrome itself to be a broader category of HES [20]. This reflects the fact that the disease concepts of HE and HES are not fully defined, and their classification has evolved over time. In any case, the patient exhibited a very advanced and dramatic clinical presentation, with no spontaneous remission for more than 6 months, leading to a diagnosis of severe Gleich syndrome after all other differential diagnoses were ruled out.

Gleich syndrome is a rare disease with no currently approved or clearly defined diagnostic criteria, and to date, fewer than 100 adult cases have been reported [21]. Nevertheless, patients with Gleich syndrome typically present with homogeneous clinical symptoms and well-characterized laboratory findings, suggesting that the disease should be considered a distinct eosinophilic disorder, likely caused by a common pathogenic mechanism [19]. In cases with clear and intense symptoms, as in this case, even a small number of disease samples are likely to lead to the identification of some pathological factors by Multi-Omics Analysis, which is very significant for the purpose of elucidating the pathology of rare diseases. This is very significant for the purpose of elucidating the pathology of rare diseases.

In the disease variant analysis using WGS data, deletion mutations associated with FOXO1, PRDM16, SPAST, ITSN1, and DLGAP2 were identified in AnnotSV as variants with an ACMG class >3 and a LOEUF_bin score of 0. The FOXO family, including FOXO1, is known to upregulate various pro-inflammatory cytokines, including interleukin (IL)- 1β, IL-9, Toll-like receptor (TLR)1, and TLR4 [22]. FOXO1 is activated by bacteria in dendritic cells (DCs) and plays crucial roles in immune responses, including promoting DC phagocytosis, migration, homing to lymph nodes, stimulating T cells, activating B cells, and enhancing antibody production [23]. The PRDM16 gene regulates adipocyte differentiation and thermogenesis, making it a promising target for the treatment of obesity and diabetes [24,25]. It is also involved in the development and function of endothelial cells and vascular smooth muscle cells in arteries and plays a critical role in the regulation of inflammatory responses [26]. ITSN1 is known to play a crucial role in pathogen uptake through clathrin-mediated endocytosis (CME) and also functions as a scaffolding protein in cell signaling pathways [27]. This may allow ITSN1 to influence eosinophil activation and function.SPAST and DLGAP2 are both genes that are critical for maintaining the health and function of the nervous system and have not been reported to be involved in eosinophil-related diseases.

SNPnexus-based analysis identified PTPRJ, SLC4A1, FGFR4, and C1GALT1C1 as genes with corresponding mutations classified as pathogenic in the ClinVar annotations. PTPRJ and FGFR4 are implicated in the development and susceptibility to various cancers [28–37]. Eosinophils are known to play an anti-tumor role in melanoma, gastric, colorectal, oral, and prostate cancers, while they are associated with poor prognosis in Hodgkin’s lymphoma and cervical cancer [38]. The relationship between eosinophilia-associated SNPs and tumor grade warrants further investigation. SLC4A1 (Solute Carrier Family 4 Member 1) encodes an anion exchange protein critical for erythrocyte function, and mutations in SLC4A1 have been shown to impair erythrocyte membrane stability [39]. However, no involvement in eosinophil-related diseases has been reported to date. C1GALT1C1 is crucial for glycosylation, and patients with Tn polyagglutination syndrome (TNPS) are known to carry an aspartic acid to glutamic acid mutation at position 131 [40].

Confirmation of mutations by exon using wANNOVAR identified a series of mutations across diverse genes, including APC, OR13C8, PCDH17, GSPT1, MED1, SLC35G4, and MADCAM1. The APC gene has been associated with familial colorectal adenomatosis, as documented on the ClinVar website [41]. PCDH17 is primarily involved in neuronal adhesion and synapse formation [42]. GSPT1 is a gene implicated in cell proliferation and translation regulation. MED1 functions as a transcriptional regulator in the nucleus, controlling the expression of specific genes [43,44]. MADCAM1, a cell adhesion molecule, is expressed in high endothelial venules, facilitating immune cell migration from the bloodstream to the gut [45]. However, no studies have been identified that directly link any of these genes to eosinophilia.

In the analysis of DEGs between patients and healthy controls from RNA-seq data, the Biological Process category of GO term enrichment analysis showed enrichment in ‘regulation of response to biotic stimulus,’ ‘response to virus,’ and GO terms related to ‘regulation of viral processes.’ Hypereosinophilia refers to an increase in the number of eosinophils in the blood, observed in response to various biotic stimuli such as infections, allergic reactions, viruses, and certain autoimmune diseases [46–48]. Eosinophils are a type of white blood cell that plays a crucial role in the body’s immune response to parasites and allergens. Their regulation and proliferation are known to be influenced by various cytokines and transcription factors, most notably interleukin-5 (IL-5) [49]. The identification of several genes enriched by biotic stimuli in this patient also reflects a pathology that results in a dramatic systemic clinical presentation.

In addition, within the Molecular Function category, several GO terms were enriched for carbohydrate binding, monosaccharide binding, glucose binding, proteoglycan binding, and glycan-related genes, such as ‘pentosyltransferase activity.’ Glycosylation is a common post-translational modification of proteins and lipids and serves as a critical recognition determinant in cell-immune system interactions. Immune and stromal cells are equipped with glycan-binding proteins (GBPs) that sense and decode diverse glycans [50,51]. This network of glycans and GBPs is crucial for the recognition of pathogens and the regulation of inflammatory and autoimmune responses, with alterations in the cellular glycome ultimately leading to pathological phenotypes [52–54]. C1GALT1C1 (rs17261572), also known as Cosmc, identified in variant analysis, galactosylates Tn antigen (GalNAcα1-Ser/Thr-R) during the biosynthesis of mucin-type O-glycans to core 1 Galβ1-3GalNAcα1-Ser/Thr (T antigen) [55] (Figure S8). Given that glycans function as drug recognition markers, the potential involvement of glycan-related genes in drug regulation has been investigated in public database analyses [56]. The relationship between rs17261572 on C1GALT1C1 and HE/Gleich syndrome, as well as its connection to other glycan-related genes, warrants further investigation.

The Z-score values derived from WGS annotation impact, along with the ranking of DEGs and PPI network analysis from RNA-seq data, identified IFI27, OAS2, and ACE as key candidate genes. In this case, the expression levels of these genes were variable compared to healthy subjects and showed a recovery trend after treatment. It is well established that a reduction in ACE leads to an increase in bradykinin, which contributes to the development of angioedema [57,58]. Bradykinin itself induces angioedema [59] and further increases phospholipase A2 activity, enhances arachidonic acid metabolism, and triggers a wide range of inflammatory responses [60]. Additionally, large amounts of bradykinin have been reported to recruit eosinophils [60,61]. In this patient, who presented with severe systemic inflammation in addition to edema, ACE was found to play a major role as a pathological factor. The patient achieved a good state of remission with oral prednisolone treatment, not only due to the general anti-inflammatory effects of corticosteroids but also because corticosteroids have been suggested to directly restore ACE expression [62], which may have contributed to the improvement in the condition.

Notably, there is a case report of a patient with congenital ACE deficiency presenting with recurrent angioedema [63]. This finding further supports the involvement of ACE as a pathological factor in diseases characterized by angioedema (Figure 3). However, in the present case, no mutation in the ACE gene itself critically defines a loss of expression, suggesting that an alteration in an upstream factor leading to reduced ACE expression may be involved.

**Figure 3.**
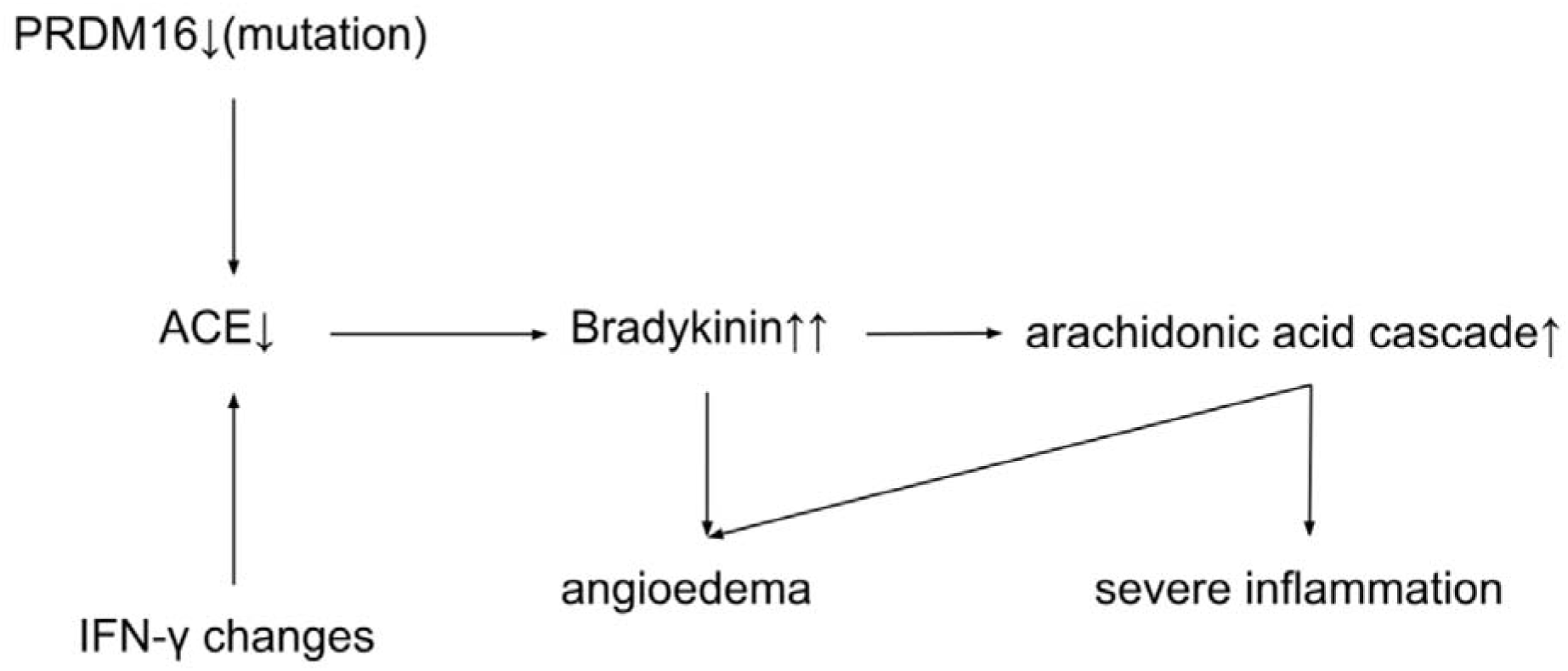
The role of ACE as a pathogenic factor in a type of HE/Gleich syndrome This figure illustrates the pathway by which a PRDM16 mutation leads to pathogenic outcomes in HE/Gleich Syndrome. The mutation downregulates ACE expression, resulting in elevated bradykinin levels. This triggers the arachidonic acid cascade, which contributes to both angioedema and severe inflammation. Additionally, IFN-γ levels influence ACE expression, further exacerbating these processes.

PRDM16, a gene with a confirmed structural abnormality in this case, has been shown to reduce ACE mRNA expression in the systemic RAAS when conditionally knocked out (CKO) in the heart, although the detailed mechanism remains unclear [64]. While ACE expression in the intrarenal RAAS is compensated for in this CKO model, it is plausible that the PRDM16 mutation observed in this patient results in a systemic reduction in ACE expression. However, the direct mechanism by which PRDM16 mutations lead to reduced ACE expression is not fully understood and warrants further investigation.

Upstream regulators and causal networks of ACE were investigated using IPA to explore their relationship with the pathogenesis in greater detail. The IFNG gene was identified as an upstream regulator of ACE. Previous studies have reported that human IFNG protein increases the expression of ACE mRNA in NB4 cells and enhances the expression of ACE protein on the cell surface of cultured human peripheral blood monocytes [65]. Additionally, IFNG has been detected in the supernatant of cultured peripheral blood mononuclear cells from patients with idiopathic eosinophilia, suggesting its involvement in the disease’s pathogenesis [66]. Further research is needed to elucidate these mechanisms in more detail.

This study has certain limitations. While Multi-Omics analyses using whole genome sequencing (WGS) and RNA sequencing (RNA-seq) have advanced our understanding of the pathogenesis of HE/Gleich syndrome at the genetic and gene expression levels, these approaches must be supplemented with protein-level analyses and in vivo model-based studies. Confirming blood levels of bradykinin and ACE would further substantiate the pathogenesis of this condition.

Moreover, other factors influencing ACE expression need to be investigated, and detailed mechanisms must be elucidated. Gene expression data alone do not adequately capture post-translational modifications, protein interactions, or functional dynamics within cells and tissues. In vivo models can provide a more comprehensive understanding of disease mechanisms within a physiological context. Therefore, future research should incorporate proteomic analyses, gene editing techniques, and animal models.

## Conclusion

The aim of this study was to analyze whole genome sequencing (WGS) and RNA sequencing (RNA-seq) data from patient-derived samples of individuals diagnosed with severe HE/Gleich syndrome of unknown etiology, and to identify the underlying pathological factors. The analysis revealed that ACE plays a significant role in the condition, demonstrating the utility of genomic analysis for understanding this disease. Specifically, the study indicated that reduced ACE expression may result from a complex interplay of factors, including PRDM16 gene mutations and altered expression of IFNγ, which may contribute to the development of angioedema and marked inflammation. The findings of this study will enhance the understanding of the pathomechanisms of HE/Gleich syndrome and inform the development of new therapeutic strategies in the future.

## Data Availability

All data produced in the present study are available upon reasonable request to the authors

## Acknowledgements

We sincerely thank the patients who participated in this study for their invaluable cooperation. I also wish to express our deepest appreciation to Dr. Hirokazu Muraoka for his extensive guidance and support throughout the entirety of this study. His contributions spanned from the conceptualization of the research, the collection of blood samples, and the acquisition of WGS and RNA-seq data, to the writing, editing, and discussion of the manuscript.

## Supplementary Information

Data Availability

## Conflict of interest statement

The authors declare that there are no conflicts.

## Notes

### Competing Interest Statement

The authors have declared no competing interest.

### Funding Statement

This study did not receive any funding

### Author Declarations

Ethics committee/IRB of CLINIC FOR Tamachi gave ethical approval for this work.

### Summary of Updates

Add Dr.sato email address to Author list.

## Reference

(1) Roufosse, F.; Weller, P. F. Journal of Allergy and Clinical Immunology 2010, 126, 39–44. doi:10.1016/j.jaci.2010.04.011

(2) Rothenberg, M. E. New England Journal of Medicine 1998, 338, 1592–1600. doi:10.1056/NEJM199805283382206

(3) Valent, P.; Klion, A. D.; Horny, H. P.; Roufosse, F.; Gotlib, J.; Weller, P. F.; Hellmann, A.; Metzgeroth, G.; Leiferman, K. M.; Arock, M.; Butterfield, J. H.; Sperr, W. R.; Sotlar, K.; Vandenberghe, P.; Haferlach, T.; Simon, H. U.; Reiter, A.; Gleich, G. J. Journal of Allergy and Clinical Immunology 2012, 130, 607–612.e9. doi:10.1016/j.jaci.2012.02.019

(4) Gleich, G. J.; Schroeter, A. L.; Marcoux, J. P.; Sachs, M. I.; O’Connell, E. J.; Kohler, P. F. Trans Assoc Am Physicians 1984, 97, 25–32. doi:10.1056/NEJM198406213102501

(5) Cools, J.; DeAngelo, D. J.; Gotlib, J.; Stover, E. H.; Legare, R. D.; Cortes, J.; Kutok, J.; Clark, J.; Galinsky, I.; Griffin, J. D.; Cross, N. C. P.; Tefferi, A.; Malone, J.; Alam, R.; Schrier, S. L.; Schmid, J.; Rose, M.; Vandenberghe, P.; Verhoef, G.; Boogaerts, M.; Wlodarska, I.; Kantarjian, H.; Marynen, P.; Coutre, S. E.; Stone, R.; Gilliland, D. G. New England Journal of Medicine 2003, 348, 1201–1214. doi:10.1056/NEJMOA025217/ASSET/02DE2273-F36A-40F0-A786-276BD835B728/ASSETS/IMAGES/LARGE/NEJMOA025217_F5.JPG

(6) Simon, H.-U.; Plötz, S. G.; Dummer, R.; Blaser, K. New England Journal of Medicine 1999, 341, 1112– 1120. doi:10.1056/NEJM199910073411503/ASSET/F1312766-756C-4DAD-BB7E-B527821BB498/ASSETS/IMAGES/LARGE/NEJM199910073411503_T4.JPG

(7) Van El, C. G.; Cornel, M. C.; Borry, P.; Hastings, R. J.; Fellmann, F.; Hodgson, S. V.; Howard, H. C.; Cambon-Thomsen, A.; Knoppers, B. M.; Meijers-Heijboer, H.; Scheffer, H.; Tranebjaerg, L.; Dondorp, W.; De Wert, G. M. W. R. European Journal of Human Genetics 2013 21:6 2013, 21, 580–584. doi:10.1038/ejhg.2013.46

(8) Ashley, E. A.; Butte, A. J.; Wheeler, M. T.; Chen, R.; Klein, T. E.; Dewey, F. E.; Dudley, J. T.; Ormond, K. E.; Pavlovic, A.; Morgan, A. A.; Pushkarev, D.; Neff, N. F.; Hudgins, L.; Gong, L.; Hodges, L. M.; Berlin, D. S.; Thorn, C. F.; Sangkuhl, K.; Hebert, J. M.; Woon, M.; Sagreiya, H.; Whaley, R.; Knowles, J. W.; Chou, M. F.; Thakuria, J. V.; Rosenbaum, A. M.; Zaranek, A. W.; Church, G. M.; Greely, H. T.; Quake, S. R.; Altman, R. B. The Lancet 2010, 375, 1525–1535. doi:10.1016/S0140-6736(10)60452-7

(9) Lupski, J. R.; Reid, J. G.; Gonzaga-Jauregui, C.; Rio Deiros, D.; Chen, D. C. Y.; Nazareth, L.; Bainbridge, M.; Dinh, H.; Jing, C.; Wheeler, D. A.; McGuire, A. L.; Zhang, F.; Stankiewicz, P.; Halperin, J. J.; Yang, C.; Gehman, C.; Guo, D.; Irikat, R. K.; Tom, W.; Fantin, N. J.; Muzny, D. M.; Gibbs, R. A. New England Journal of Medicine 2010, 362, 1181–1191. doi:10.1056/NEJMOA0908094/SUPPL_FILE/NEJMOA0908094_DISCLOSURES.PDF

(10) Roach, J. C.; Glusman, G.; Smit, A. F. A.; Huff, C. D.; Hubley, R.; Shannon, P. T.; Rowen, L.; Pant, K. P.; Goodman, N.; Bamshad, M.; Shendure, J.; Drmanac, R.; Jorde, L. B.; Hood, L.; Galas, D. J. Science (1979) 2010, 328, 636–639. doi:10.1126/SCIENCE.1186802/SUPPL_FILE/ROACH-SOM.PDF

(11) Lee, J. S.; Seo, H.; Im, K.; Park, S. N.; Kim, S. M.; Lee, E. K.; Kim, J. A.; Lee, J. H.; Kwon, S.; Kim, M.; Koh, I.; Hwang, S.; Park, H. W.; Kang, H. R.; Park, K. S.; Kim, J. H.; Lee, D. S. PLoS One 2017, 12, e0185602. doi:10.1371/JOURNAL.PONE.0185602

(12) Andersen, C. L.; Nielsen, H. M.; Kristensen, L. S.; Søgaard, A.; Vikeså, J.; Jønson, L.; Nielsen, F. C.; Hasselbalch, H.; Bjerrum, O. W.; Punj, V.; Grønbæk, K.; Andersen, C. L.; Nielsen, H. M.; Kristensen, L. S.; Søgaard, A.; Vikeså, J.; Jønson, L.; Nielsen, F. C.; Hasselbalch, H.; Bjerrum, O. W.; Punj, V.; Grønbæk, K. Oncotarget 2015, 6, 40588–40597. doi:10.18632/ONCOTARGET.5845

(13) Blake, J. A.; Christie, K. R.; Dolan, M. E.; Drabkin, H. J.; Hill, D. P.; Ni, L.; Sitnikov, D.; Burgess, S.; Buza, T.; Gresham, C.; McCarthy, F.; Pillai, L.; Wang, H.; Carbon, S.; Dietze, H.; Lewis, S. E.; Mungall, C. J.; Munoz-Torres, M. C.; Feuermann, M.; Gaudet, P.; Basu, S.; Chisholm, R. L.; Dodson, R. J.; Fey, P.; Mi, H.; Thomas, P. D.; Muruganujan, A.; Poudel, S.; Hu, J. C.; Aleksander, S. A.; McIntosh, B. K.; Renfro, D. P.; Siegele, D. A.; Attrill, H.; Brown, N. H.; Tweedie, S.; Lomax, J.; Osumi-Sutherland, D.; Parkinson, H.; Roncaglia, P.; Lovering, R. C.; Talmud, P. J.; Humphries, S. E.; Denny, P.; Campbell, N. H.; Foulger, R. E.; Chibucos, M. C.; Giglio, M. G.; Chang, H. Y.; Finn, R.; Fraser, M.; Mitchell, A.; Nuka, G.; Pesseat, S.; Sangrador, A.; Scheremetjew, M.; Young, S. Y.; Stephan, R.; Harris, M. A.; Oliver, S. G.; Rutherford, K.; Wood, V.; Bahler, J.; Lock, A.; Kersey, P. J.; McDowall, M. D.; Staines, D. M.; Dwinell, M.; Shimoyama, M.; Laulederkind, S.; Hayman, G. T.; Wang, S. J.; Petri, V.; D’Eustachio, P.; Matthews, L.; Balakrishnan, R.; Binkley, G.; Cherry, J. M.; Costanzo, M. C.; Demeter, J.; Dwight, S. S.; Engel, S. R.; Hitz, B. C.; Inglis, D. O.; Lloyd, P.; Miyasato, S. R.; Paskov, K.; Roe, G.; Simison, M.; Nash, R. S.; Skrzypek, M. S.; Weng, S.; Wong, E. D.; Berardini, T. Z.; Li, D.; Huala, E.; Argasinska, J.; Arighi, C.; Auchincloss, A.; Axelsen, K.; Argoud-Puy, G.; Bateman, A.; Bely, B.; Blatter, M. C.; Bonilla, C.; Bougueleret, L.; Boutet, E.; Breuza, L.; Bridge, A.; Britto, R.; Casals, C.; Cibrian-Uhalte, E.; Coudert, E.; Cusin, I.; Duek-Roggli, P.; Estreicher, A.; Famiglietti, L.; Gane, P.; Garmiri, P.; Gos, A.; Gruaz-Gumowski, N.; Hatton-Ellis, E.; Hinz, U.; Hulo, C.; Huntley, R.; Jungo, F.; Keller, G.; Laiho, K.; Lemercier, P.; Lieberherr, D.; Macdougall, A.; Magrane, M.; Martin, M.; Masson, P.; Mutowo, P.; O’Donovan, C.; Pedruzzi, I.; Pichler, K.; Poggioli, D.; Poux, S.; Rivoire, C.; Roechert, B.; Sawford, T.; Schneider, M.; Shypitsyna, A.; Stutz, A.; Sundaram, S.; Tognolli, M.; Wu, C.; Xenarios, I.; Chan, J.; Kishore, R.; Sternberg, P. W.; Van Auken, K.; Muller, H. M.; Done, J.; Li, Y.; Howe, D.; Westerfeld, M. Nucleic Acids Res 2015, 43, D1049–D1056. doi:10.1093/NAR/GKU1179

(14) Ghebrehiwet, B.; Joseph, K.; Kaplan, A. P. Frontiers in Allergy 2024, 5, 1302605. doi:10.3389/FALGY.2024.1302605/BIBTEX

(15) Straka, B. T.; Ramirez, C. E.; Byrd, J. B.; Stone, E.; Woodard-Grice, A.; Nian, H.; Yu, C.; Banerji, A.; Brown, N. J. Journal of Allergy and Clinical Immunology 2017, 140, 242–248.e2. doi:10.1016/j.jaci.2016.09.051

(16) Simon, H.-U.; Plötz, S. G.; Dummer, R.; Blaser, K. New England Journal of Medicine 1999, 341, 1112– 1120. doi:10.1056/NEJM199910073411503/ASSET/F1312766-756C-4DAD-BB7E-B527821BB498/ASSETS/IMAGES/LARGE/NEJM199910073411503_T4.JPG

(17) Basso, J. R.; Bizinoto, L. G. Z.; Limone, G. A.; Enokihara, M. M. S. S.; Do Espirito-Santo Filho, K.; Fonseca, A. R.; Agondi, R. C.; de Gois, A. F. T.; Cunha, L. L. Brazilian Journal of Medical and Biological Research 2021, 54, e10745. doi:10.1590/1414-431X202010745

(18) Mormile, I.; Petraroli, A.; Loffredo, S.; Rossi, F. W.; Mormile, M.; Mastro, A. Del; Spadaro, G.; de Paulis, A.; Bova, M. Journal of Clinical Medicine 2021, Vol. 10, *Page* 1442 2021, *10*, 1442. doi:10.3390/JCM10071442

(19) Khoury, P.; Herold, A.; Alpaugh, A.; Dinerman, E.; Holland-Thomas, N.; Stoddard, J.; Gurprasad, S.; Maric, I.; Simakova, O.; Schwartz, L. B.; Fong, J.; Richard Lee, C. C.; Xi, L.; Wang, Z.; Raffeld, M.; Klion, A. D. Haematologica 2015, 100, 300–307. doi:10.3324/HAEMATOL.2013.091264

(20) Valent, P.; Klion, A. D.; Horny, H. P.; Roufosse, F.; Gotlib, J.; Weller, P. F.; Hellmann, A.; Metzgeroth, G.; Leiferman, K. M.; Arock, M.; Butterfield, J. H.; Sperr, W. R.; Sotlar, K.; Vandenberghe, P.; Haferlach, T.; Simon, H. U.; Reiter, A.; Gleich, G. J. Journal of Allergy and Clinical Immunology 2012, 130, 607–612.e9. doi:10.1016/j.jaci.2012.02.019

(21) Mormile, I.; Petraroli, A.; Loffredo, S.; Rossi, F. W.; Mormile, M.; Mastro, A. Del; Spadaro, G.; de Paulis, A.; Bova, M. Journal of Clinical Medicine 2021, Vol. 10, *Page* 1442 2021, *10*, 1442. doi:10.3390/JCM10071442

(22) Kim, M. E.; Kim, D. H.; Lee, J. S. International Journal of Molecular Sciences 2022, *Vol.* 23, *Page 11877* 2022, *23*, 11877. doi:10.3390/IJMS231911877

(23) Cabrera-Ortega, A. A.; Feinberg, D.; Liang, Y.; Rossa, C.; Graves, D. T. *Critical Reviews&*trade; in Immunology 2017, 37, 1–13. doi:10.1615/CRITREVIMMUNOL.2017019636

(24) Seale, P.; Conroe, H. M.; Estall, J.; Kajimura, S.; Frontini, A.; Ishibashi, J.; Cohen, P.; Cinti, S.; Spiegelman, B. M. J Clin Invest 2011, 121, 96–105. doi:10.1172/JCI44271

(25) Jiang, N.; Yang, M.; Han, Y.; Zhao, H.; Sun, L. Front Pharmacol 2022, 13, 870250. doi:10.3389/FPHAR.2022.870250/BIBTEX

(26) Thompson, M.; Sakabe, M.; Verba, M.; Hao, J.; Meadows, S. M.; Lu, Q. R.; Xin, M. Front Physiol 2023, 14, 1165379. doi:10.3389/FPHYS.2023.1165379/BIBTEX

(27) Gerasymchuk, D.; Hubiernatorova, A.; Domanskyi, A. Front Neurol 2020, 11, 549006. doi:10.3389/FNEUR.2020.549006/BIBTEX

(28) Iuliano, R.; Le Pera, I.; Cristofaro, C.; Baudi, F.; Arturi, F.; Pallante, P. L.; Martelli, M. L.; Trapasso, F.; Chiariotti, L.; Fusco, A. Oncogene 2004 23:52 2004, 23, 8432–8438. doi:10.1038/sj.onc.1207766

(29) Jaakkola, S.; Salmikangas, P.; Nylund, S.; Lehtovirta, P.; Nevanlinna, H.; Partanen, J.; Armstrong, E.; Pyrhönen, S. Int J Cancer 1993, 54, 378–382. doi:10.1002/IJC.2910540305

(30) Leung, H. Y.; Gullick, W. J.; Lemoine, N. R. Int J Cancer 1994, 59, 667–675. doi:10.1002/IJC.2910590515

(31) Sahadevan, K.; Darby, S.; Leung, H. Y.; Mathers, M. E.; Robson, C. N.; Gnanapragasam, V. J. J Pathol 2007, 213, 82–90. doi:10.1002/PATH.2205

(32) Streit, S.; Mestel, D. S.; Schmidt, M.; Ullrich, A.; Berking, C. British Journal of Cancer 2006 94:12 2006, 94, 1879–1886. doi:10.1038/sj.bjc.6603181

(33) Cancer Progression and Tumor Cell Motility Are Associated with the FGFR4 Arg388 Allele | Cancer Research | American Association for Cancer Research https://aacrjournals.org/cancerres/article/62/3/840/509605/Cancer-Progression-and-Tumor-Cell-Motility-Are (accessed Aug 16, 2024)

(34) Matakidou, A.; El Galta, R.; Rudd, M. F.; Webb, E. L.; Bridle, H.; Eisen, T.; Houlston, R. S. British Journal of Cancer 2007 96:12 2007, 96, 1904–1907. doi:10.1038/sj.bjc.6603816

(35) Spinola, M.; Leani, V.; Pignatiello, C.; Conti, B.; Ravagnani, F.; Pastorino, U.; Dragani, T. A. 0.1200/JCO.2005.17.350 2016, *23*, 7307–7311. doi:10.1200/JCO.2005.17.350

(36) Wang, J.; Stockton, D. W.; Ittmann, M. Clinical Cancer Research 2004, 10, 6169–6178. doi:10.1158/1078-0432.CCR-04-0408

(37) Thussbas, C.; Nahrig, J.; Streit, S.; Bange, J.; Kriner, M.; Kates, R.; Ulm, K.; Kiechle, M.; Hoefler, H.; Ullrich, A.; Harbeck, N. 10.1200/JCO.2005.04.8587 2016, *24*, 3747–3755. doi:10.1200/JCO.2005.04.8587

(38) Varricchi, G.; Galdiero, M. R.; Loffredo, S.; Lucarini, V.; Marone, G.; Mattei, F.; Marone, G.; Schiavoni, G. Oncoimmunology 2018, 7. doi:10.1080/2162402X.2017.1393134

(39) Bruce, L. J.; Robinson, H. C.; Guizouarn, H.; Borgese, F.; Harrison, P.; King, M. J.; Goede, J. S.; Coles, S. E.; Gore, D. M.; Lutz, H. U.; Ficarella, R.; Layton, D. M.; Iolascon, A.; Ellory, J. C.; Stewart, G. W. Nature Genetics 2005 37:11 2005, 37, 1258–1263. doi:10.1038/ng1656

(40) Ju, T.; Cummings, R. D. Proc Natl Acad Sci U S A 2002, 99, 16613–16618. doi:10.1073/PNAS.262438199/ASSET/E7D75E83-13F7-4461-A8AA-B2B085A2E14B/ASSETS/GRAPHIC/PQ2624381004.JPEG

(41) VCV001050412.3 - ClinVar - NCBI https://www.ncbi.nlm.nih.gov/clinvar/variation/1050412/ (accessed Aug 16, 2024)

(42) Abrahams, B. S.; Tentler, D.; Perederiy, J. V.; Oldham, M. C.; Coppola, G.; Geschwind, D. H. Proc Natl Acad Sci U S A 2007, 104, 17849–17854. doi:10.1073/PNAS.0706128104/SUPPL_FILE/06128TABLE6.PDF

(43) Hoshino, S.; Imai, M.; Mizutani, M.; Kikuchi, Y.; Hanaoka, F.; Ui, M.; Katada, T. Journal of Biological Chemistry 1998, 273, 22254–22259. doi:10.1074/jbc.273.35.22254

(44) Kikuchi, Y.; Shimatake, H.; Kikuchi, A. EMBO J 1988, 7, 1175–1182. doi:10.1002/J.1460-2075.1988.TB02928.X

(45) Arihiro, S.; Ohtani, H.; Suzuki, M.; Murata, M.; Ejima, C.; Oki, M.; Kinouchi, Y.; Fukushima, K.; Sasaki, I.; Nakamura, S.; Matsumoto, T.; Torii, A.; Toda, G.; Nagura, H. Pathol Int 2002, 52, 367–374. doi:10.1046/J.1440-1827.2002.01365.X

(46) Macchia, I.; La Sorsa, V.; Urbani, F.; Moretti, S.; Antonucci, C.; Afferni, C.; Schiavoni, G. Front Immunol 2023, 14, 1170035. doi:10.3389/FIMMU.2023.1170035/BIBTEX

(47) Thomsen, G. N.; Christoffersen, M. N.; Lindegaard, H. M.; Davidsen, J. R.; Hartmeyer, G. N.; Assing, K.; Mortz, C. G.; Martin-Iguacel, R.; Møller, M. B.; Kjeldsen, A. D.; Havelund, T.; El Fassi, D.; Broesby-Olsen, S.; Maiborg, M.; Johansson, S. L.; Andersen, C. L.; Vestergaard, H.; Bjerrum, O. W. Front Oncol 2023, 13, 1193730. doi:10.3389/FONC.2023.1193730/BIBTEX

(48) Klion, A. D. Hematology 2015, 2015, 92–97. doi:10.1182/ASHEDUCATION-2015.1.92

(49) Kouro, T.; Takatsu, K. Int Immunol 2009, 21, 1303–1309. doi:10.1093/INTIMM/DXP102

(50) Alves, I.; Fernandes, Â.; Santos-Pereira, B.; Azevedo, C. M.; Pinho, S. S. FEBS Lett 2022, 596, 1485–1502. doi:10.1002/1873-3468.14347

(51) Pinho, S. S.; Alves, I.; Gaifem, J.; Rabinovich, G. A. Cellular & Molecular Immunology 2023 *20:10* 2023, 20, 1101–1113. doi:10.1038/s41423-023-01074-1

(52) Ohtsubo, K.; Marth, J. D. Cell 2006, 126, 855–867. doi:10.1016/J.CELL.2006.08.019

(53) Pinho, S. S.; Reis, C. A. Nature Reviews Cancer 2015 15:9 2015, 15, 540–555. doi:10.1038/nrc3982

(54) Rabinovich, G. A.; Croci, D. O. Immunity 2012, 36, 322–335. doi:10.1016/J.IMMUNI.2012.03.004

(55) Wang, Y.; Ju, T.; Ding, X.; Xia, B.; Wang, W.; Xia, L.; He, M.; Cummings, R. D. Proc Natl Acad Sci U S A 2010, 107, 9228–9233. doi:10.1073/PNAS.0914004107/SUPPL_FILE/SFIG05.TIF

(56) Koreeda, T.; Honda, H. Glycoconj J 2024, 41, 133–149. doi:10.1007/S10719-024-10153-Y/METRICS

(57) Ghebrehiwet, B.; Joseph, K.; Kaplan, A. P. Frontiers in Allergy 2024, 5, 1302605. doi:10.3389/FALGY.2024.1302605/BIBTEX

(58) Straka, B. T.; Ramirez, C. E.; Byrd, J. B.; Stone, E.; Woodard-Grice, A.; Nian, H.; Yu, C.; Banerji, A.; Brown, N. J. Journal of Allergy and Clinical Immunology 2017, 140, 242–248.e2. doi:10.1016/j.jaci.2016.09.051

(59) Cicardi, M.; Zuraw, B. L. J Allergy Clin Immunol Pract 2018, 6, 1132–1141. doi:10.1016/J.JAIP.2018.04.022

(60) Kaplan, A. P.; Joseph, K.; Silverberg, M. Journal of Allergy and Clinical Immunology 2002, 109, 195–209. doi:10.1067/MAI.2002.121316

(61) Roisman, G. L.; Lacronique, J. G.; Desmazes-Dufeu, N.; Carré, C.; Le Cae, A.; Dusser, D. J. 10.1164/ajrccm.153.1.8542147 2012, *153*, 381–390. doi:10.1164/AJRCCM.153.1.8542147

(62) Fishel, R. S.; Eisenberg, S.; Shai, S. Y.; Redden, R. A.; Bernstein, K. E.; Berk, B. C. Hypertension 1995, 25, 343–349. doi:10.1161/01.HYP.25.3.343/ASSET/9C5DA6FC-13D9-494B-97A2-3A87C146FD08/ASSETS/GRAPHIC/HY0350384008.JPEG

(63) Sreeram, A. B.; Corey, J. P. 10.1016/S0194-59989570277-6 1995, 112, 421–423. doi:10.1016/S0194-59989570277-6

(64) Kang, J. O.; Ha, T. W.; Jung, H. U.; Lim, J. E.; Oh, B. PLoS One 2022, 17, e0267938. doi:10.1371/JOURNAL.PONE.0267938

(65) Obeid, D.; Nguyen, J.; Lesavre, P.; Bauvois, B. Oncogene 2007 26:1 2006, 26, 102–110. doi:10.1038/sj.onc.1209779

(66) Simon, H.-U.; Plötz, S. G.; Dummer, R.; Blaser, K. New England Journal of Medicine 1999, 341, 1112– 1120. doi:10.1056/NEJM199910073411503/ASSET/F1312766-756C-4DAD-BB7E-B527821BB498/ASSETS/IMAGES/LARGE/NEJM199910073411503_T4.JPG

