## Supplementary Information 2 for "A newly identified pathology of Episodic Angioedema with Hypereosinophilia (Gleich’s Syndrome) revealed by Multi-Omics Analysis"

| **Item** | **Value** | **Reference Range** |
| --- | --- | --- |
| TP | 7.6 g/dL | 6.7–8.3 g/dL |
| Albumin | 51.4% | 55.8–66.1% |
| α1-globulin | 5.2% | 2.9–4.9% |
| α2-globulin | 11.3% | 7.1–11.8% |
| β1-globulin | 4.7% | 4.7–7.2% |
| β2-globulin | 4.2% | 3.2–6.5% |
| γ-globulin | 23.2% | 11.1–18.8% |
| A/G ratio | 1.1 | 1.3–1.9 |
| T-bil | 1.1 mg/dL | 0.3–1.2 mg/dL |
| Indirect bilirubin | 0.9 mg/dL | ≤0.8 mg/dL |
| AST | 31 U/L | 10–40 U/L |
| ALT | 22 U/L | 5–40 U/L |
| LD | 328 U/L | 124–222 U/L |
| γ-GT | 14 U/L | ≤30 U/L (female) |
| HDL | 51 mg/dL | 40–96 mg/dL (female) |
| LDL | 198 mg/dL | 70–139 mg/dL |
| TG | 232 mg/dL | 50–149 mg/dL |
| UA | 4.6 mg/dL | 2.5–7.0 mg/dL (female) |
| CK | 38 U/L | 45–163 U/L (female) |
| CK-MB | <1.0 ng/mL | ≤5.0 ng/mL |
| BUN | 14.3 mg/dL | 8.0–22.0 mg/dL |
| Cr | 0.57 mg/dL | 0.47–0.79 mg/dL |
| Na | 140 mEq/L | 136–147 mEq/L |
| Cl | 101 mEq/L | 98–109 mEq/L |
| K | 3.3 mEq/L | 3.6–5.0 mEq/L |
| Ca | 8.7 mg/dL | 8.5–10.2 mg/dL |
| Serum Copper | 119 µg/dL | 68–128 µg/dL |
| WBC | 19200 /μL | 3500–9100 /μL (female) |
| RBC | 388 *10^4/μL | 376–500 *10^4/μL (female) |
| Hb | 11.1 g/dL | 11.3–15.2 g/dL (female) |
| Ht | 36.5% | 33.4–44.9% (female) |
| MCV | 94.1 fL | 79.0–100.0 fL (female) |
| MCH | 28.6 pg | 26.3–34.3 pg (female) |
| MCHC | 30.4% | 30.7–36.6% (female) |
| PLT | 28.8 *10^4/μL | 13.0–36.9 *10^4/μL (female) |
| ACTH | 3.0 pg/mL | 7.2–63.3 pg/mL |
| Cortisol | 3.8 μg/dL | 7.07–19.6 μg/dL |
| TSH | 13.5 μIU/mL | 0.61–4.23 μIU/mL |
| FreeT3 | 2.37 pg/mL | 2.52–4.06 pg/mL |
| FreeT4 | 0.88 ng/dL | 0.75–1.45 ng/dL |
| Anti-thyroglobulin antibody | 11.7 IU/mL | <19.3 IU/mL |
| Anti-TPO antibodies | 9.7 IU/mL | <3.3 IU/mL |
| CRP | 17.2 mg/dL | ≤0.14 mg/dL |
| β2-Microglobulin | 2.97 mg/dL | 1.0–1.9 mg/L |
| sIL-2R | 1668 U/mL | 157–474 U/mL |
| TARC | 291 pg/mL | <450 pg/mL (adults) |
| IgE | 504 IU/mL | ≤170 IU/mL |
| IgG | 1384 mg/dL | 870–1700 mg/dL |
| IgA | 106 mg/dL | 110–410 mg/dL |
| IgM | 136 mg/dL | 46–260 mg/dL (female) |
| IgG4 | 33 mg/dL | 11–121 mg/dL |
| ANA | <1:40 | <1:40 |
| Serum complement CH50 | 33.6 CH50/mL | 25.0–48.0 CH50/mL |
| C3 | 114 mg/dL | 86–160 mg/dL |
| C4 | 22 mg/dL | 17–45 mg/dL |
| MPO-ANCA | <1.0 U/mL | <3.5 U/mL |
| PR3-ANCA | <1.0 U/mL | <3.5 U/mL |
| Aspergillus IgG | <1.4 AU/mL | negative |
| EB Virus DNA Quantification | Not detected | Not detected (Log IU/mL) |
| HHV-6 DNA Qualitative | Negative | Negative |
| Cytomegalovirus (CF method) | 16x | <1:4 |
| Cytomegalovirus pp65 antigen (C10.C11) | Positive cell count 0/0 | Negative |

**Supplementary Table 1: General Blood Test**

| **Item** | **Value** | **Reference Range** |
| --- | --- | --- |
| Neut | 66.0% | 40.0–74.0% |
| Eosin | 16.4% | 0.0–6.0% |
| Baso | 0.3% | 0.0–2.0% |
| Mono | 1.3% | 0.0–8.0% |
| Lym | 16.0% | 18.0–59.0% |

**Supplementary Table 2: Peripheral Blood Test**

| **Item** | **Value** | **Class** |
| --- | --- | --- |
| House Dust | index 0.93 | Class 2 |
| Dust Mites | index 0.98 | Class 2 |
| Cedar Pollen | index 2.47 | Class 3 |
| Cypress Pollen | <0.27 | Class 0 |
| Alder Pollen | <0.27 | Class 0 |
| Birch Pollen | <0.27 | Class 0 |
| Timothy Grass Pollen | <0.27 | Class 0 |
| Orchard Grass Pollen | <0.27 | Class 0 |
| Ragweed Pollen | <0.27 | Class 0 |
| Mugwort Pollen | <0.27 | Class 0 |
| Alternaria Mold | <0.27 | Class 0 |
| Aspergillus | <0.27 | Class 0 |
| Candida | <0.27 | Class 0 |
| Malassezia | <0.27 | Class 0 |
| Cat (dander) | index 0.29 | Class 0 |
| Dog (dander) | <0.27 | Class 0 |
| Cockroach | <0.27 | Class 0 |
| Moth | <0.27 | Class 0 |
| Milk | <0.27 | Class 0 |
| Egg White | <0.27 | Class 0 |
| Ovomucoid | <0.27 | Class 0 |
| Rice | <0.27 | Class 0 |
| Wheat (Food) | <0.27 | Class 0 |
| Buckwheat | <0.27 | Class 0 |
| Soy | <0.27 | Class 0 |
| Peanut | <0.27 | Class 0 |
| Apple | <0.27 | Class 0 |
| Banana | <0.27 | Class 0 |
| Kiwi | <0.27 | Class 0 |
| Sesame | <0.27 | Class 0 |
| Beef | <0.27 | Class 0 |
| Pork | <0.27 | Class 0 |
| Chicken | <0.27 | Class 0 |
| Shrimp | <0.27 | Class 0 |
| Crab | <0.27 | Class 0 |
| Mackerel | <0.27 | Class 0 |
| Salmon | <0.27 | Class 0 |
| Tuna | <0.27 | Class 0 |
| Latex | <0.27 | Class 0 |

**Supplementary Table 3: Allergy Tests**

| **Item** | **Value** | **Reference Range** |
| --- | --- | --- |
| FIP1L1-PDGFRα Fusion Gene Test | Negative | Negative |
| PDGFB Split Signal | Negative | Negative |
| FGFR1 Split Signal | Negative | Negative |
| T-cell Receptor β-chain Cβ1 Reconfiguration | Negative | Negative |
| MEFV Gene Mutation | Negative | Negative |

**Supplementary Table 4: Genetic Testing**

| **Item** | **Value** | **Reference Range** |
| --- | --- | --- |
| Dog Heartworm | Class 1 | Class 0 |
| Dog Roundworm | Class 0 | Class 0 |
| Brazilian Roundworm | Class 0 | Class 0 |
| Anisakis | Class 0 | Class 0 |
| Gnathostoma (Jawed Worm) | Class 0 | Class 0 |
| Strongyloides | Class 0 | Class 0 |
| Paragonimus westermani (Lung Fluke) | Class 0 | Class 0 |
| Miyazaki Lung Fluke | Class 0 | Class 0 |
| Liver Fluke | Class 0 | Class 0 |
| Clonorchis | Class 0 | Class 0 |
| Manson's Isolated Worm | Class 0 | Class 0 |
| Taenia solium (Pork Tapeworm Cyst) | Class 0 | Class 0 |

**Supplementary Table 5: Anti-Parasitic Antibody Screening Test**

**Multimodal Diagnostic Evaluation Combining Imaging, Biopsy and Histopathological Findings**

**[CT]**
No clear abnormalities in the lungs. Pleural effusion is not evident.
Enlarged lymph nodes are noted in the axilla.
Splenomegaly is present. No other obvious abnormalities are observed in the abdominal organs.
No significant abnormalities in the pelvic region. No ascites.
No apparent abnormalities in the head and neck regions.

**Diagnosis**: No known source of fever. Prominent axillary lymph nodes.

**[MRI]**
STIR high signal changes are observed in muscles, primarily around the bilateral shoulders, including the bilateral pectoralis major, deltoid, and infraspinatus tendons.
The bilateral neck and submandibular lymph nodes are slightly enlarged, with suspected reactive enlargement.
Small bilateral pleural effusions are present.

**Diagnosis**: Suspected myositis, mainly around both shoulders.

**Needle biopsy specimen from left axillary lymph node**
**Histopathological Diagnosis**: Dermatopathic lymphadenopathy, compatible.

**Histopathological Findings**:
Lymph node tissue was obtained histologically. Lymph follicles with germinal centers and enlarged interfollicular areas are observed.
The interfollicular area contains vessels with enlarged endothelial cells, eosinophils, histiocytes, and cells with irregularly shaped nuclei and abundant mitotic activity.
These cells form clusters, creating bright areas.
Melanin deposition is prominent. No significant atypia is observed in the lymphocytes.

**Immunohistochemistry**:

- **CD20**: Positive, mainly in lymph follicles.
- **CD3, CD5**: Similar distribution, mainly positive in the interfollicular area.
- **CD10**: Positive, primarily in germinal centers.
- **Bcl-2**: Negative in the germinal centers, positive elsewhere.
- **CD4, CD8**: Predominantly positive for CD4.
- **CD21**: Mainly positive in the germinal centers, forming an FDC meshwork.
- **κ/λ**: No predominant bias.
- **Cyclin D1**: Negative, with few scattered positive cells.
- **CD1a, langerin, S-100**: Positive in bright areas.
- **MIB-1**: Positive cells consistent with germinal centers; other scattered positivity.
- **EBER (ISH)**: Negative.

**Diagnosis**: The findings are suggestive of dermatopathic lymphadenopathy. No evidence of lymphoma or other malignant features is present.

**Skin biopsy specimen**
**Histopathological Diagnosis**: Dermal perivascular inflammation.

**Histopathological Findings**:
Mild mononuclear cell infiltration is observed, primarily around the blood vessels in the dermis beneath the epidermis.
There is a mild infiltration of neutrophils and occasional nuclear debris.
Vasculitis cannot be entirely ruled out.
