## Supplementary Information 1 for "A newly identified pathology of Episodic Angioedema with Hypereosinophilia (Gleich’s Syndrome) revealed by Multi-Omics Analysis"

### Slide 1
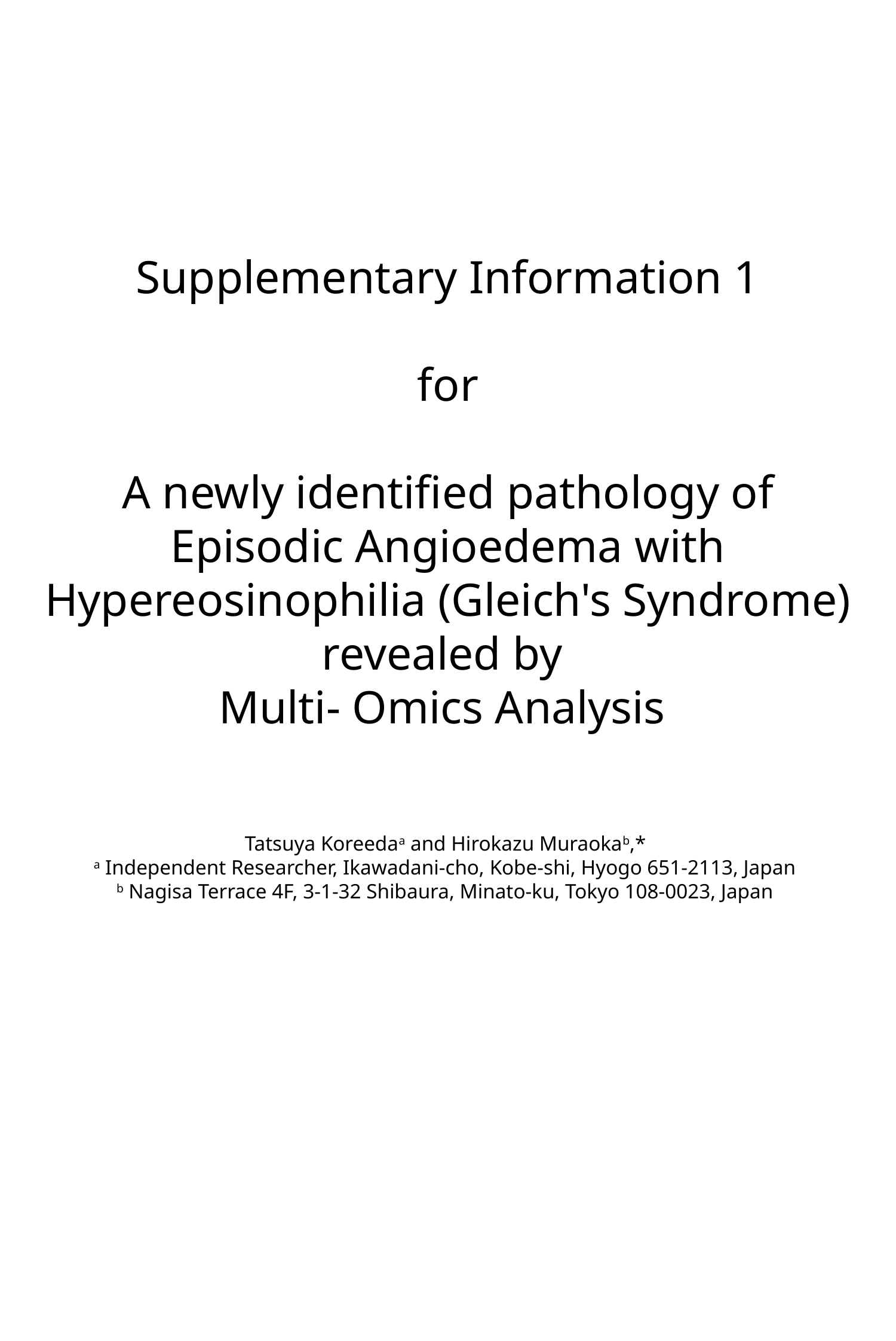

#### Supplementary Information 1 for A newly identified pathology of Episodic Angioedema with Hypereosinophilia (Gleich's Syndrome) revealed by Multi- Omics Analysis   Tatsuya Koreeda^a^ and Hirokazu Muraoka^b^,* ^a^ Independent Researcher, Ikawadani-cho, Kobe-shi, Hyogo 651-2113, Japan ^b^ Nagisa Terrace 4F, 3-1-32 Shibaura, Minato-ku, Tokyo 108-0023, Japan

### Slide 2
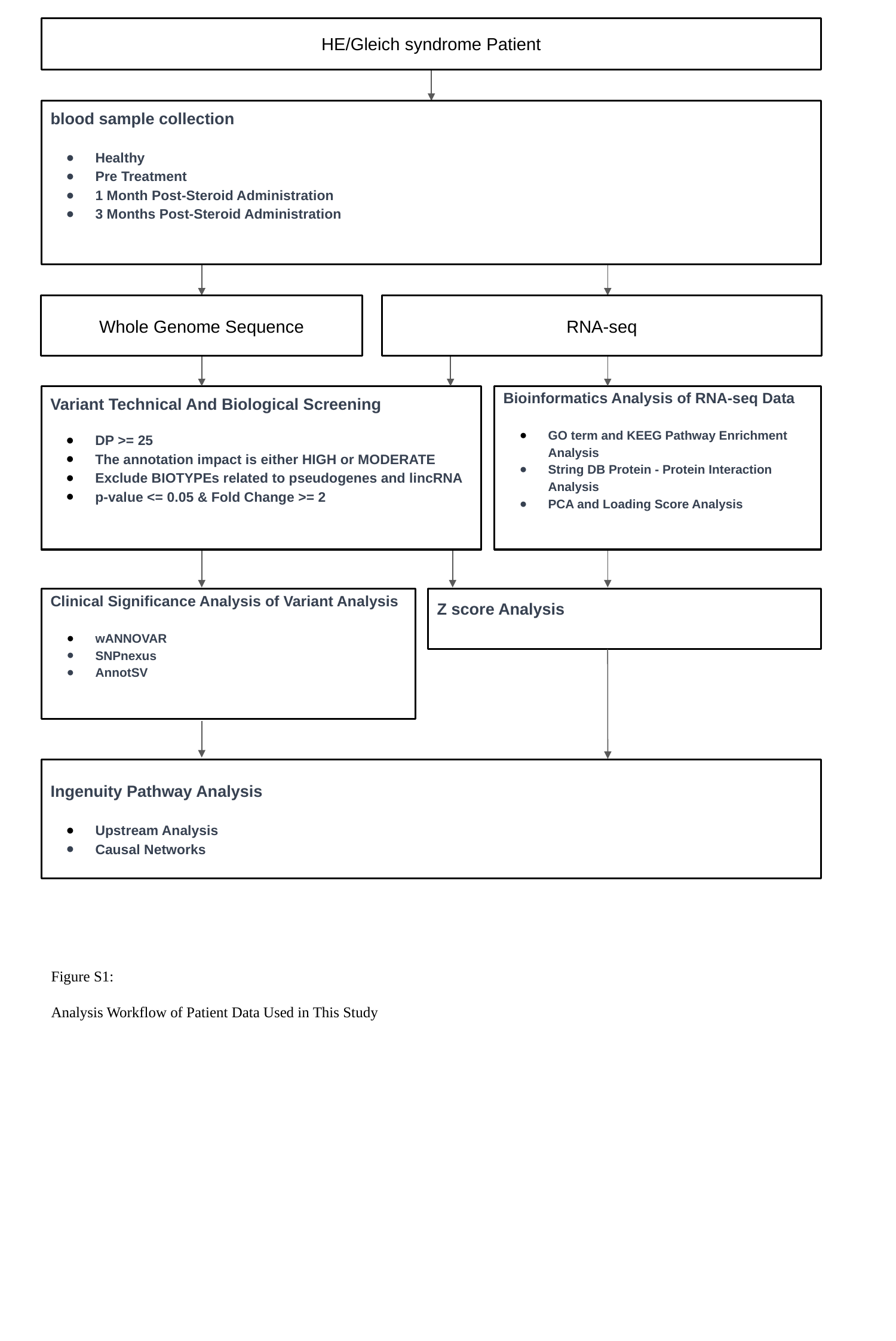

HE/Gleich syndrome Patient
blood sample collection
Healthy
Pre Treatment
1 Month Post-Steroid Administration
3 Months Post-Steroid Administration
Whole Genome Sequence
RNA-seq
Variant Technical And Biological Screening
DP >= 25
The annotation impact is either HIGH or MODERATE
Exclude BIOTYPEs related to pseudogenes and lincRNA
p-value <= 0.05 & Fold Change >= 2
Bioinformatics Analysis of RNA-seq Data
GO term and KEEG Pathway Enrichment Analysis
String DB Protein - Protein Interaction Analysis
PCA and Loading Score Analysis
Clinical Significance Analysis of Variant Analysis
wANNOVAR
SNPnexus
AnnotSV
Z score Analysis
Ingenuity Pathway Analysis
Upstream Analysis
Causal Networks
Figure S1:
Analysis Workflow of Patient Data Used in This Study

### Slide 3
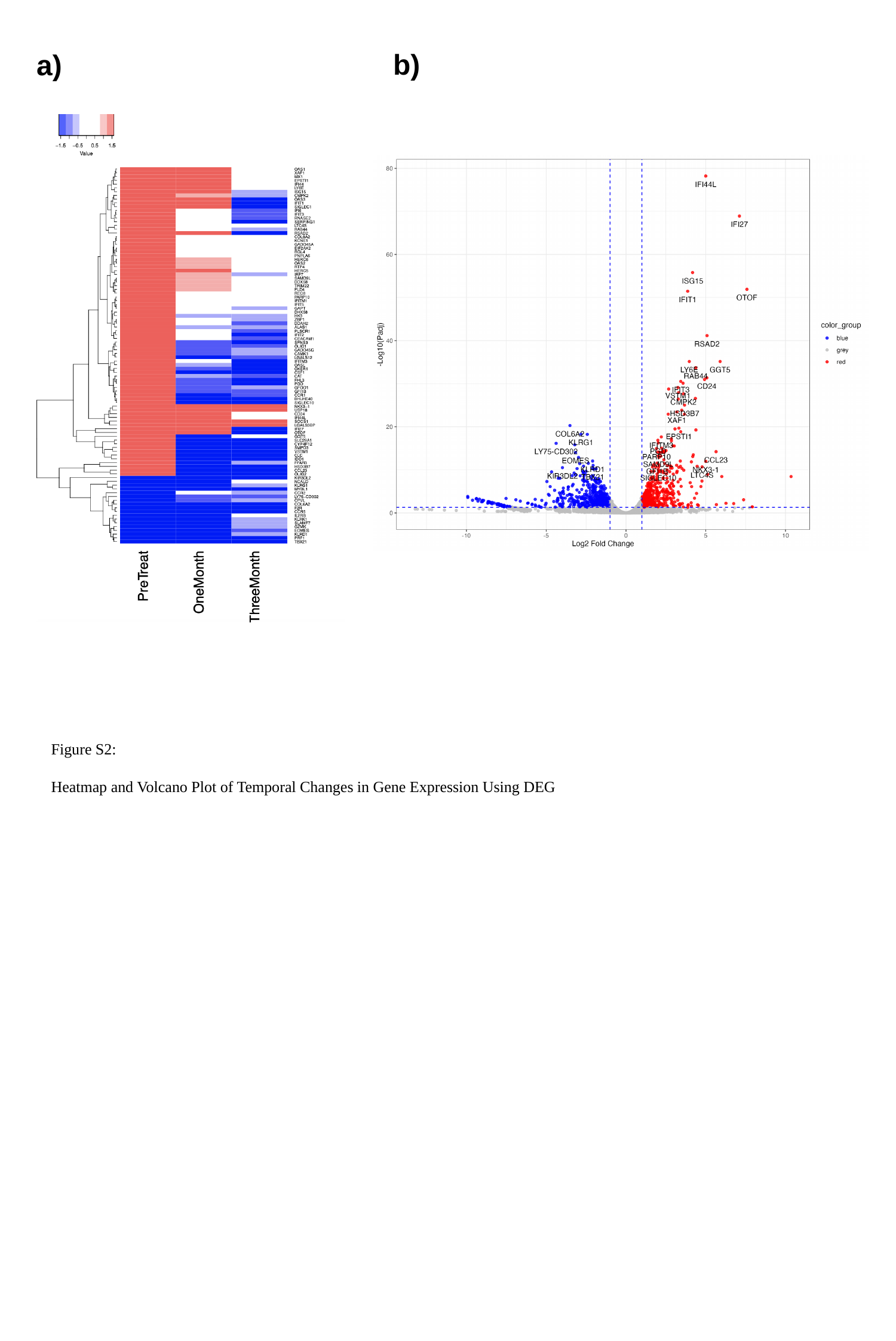

b)
a)
Figure S2:
Heatmap and Volcano Plot of Temporal Changes in Gene Expression Using DEG

### Slide 4
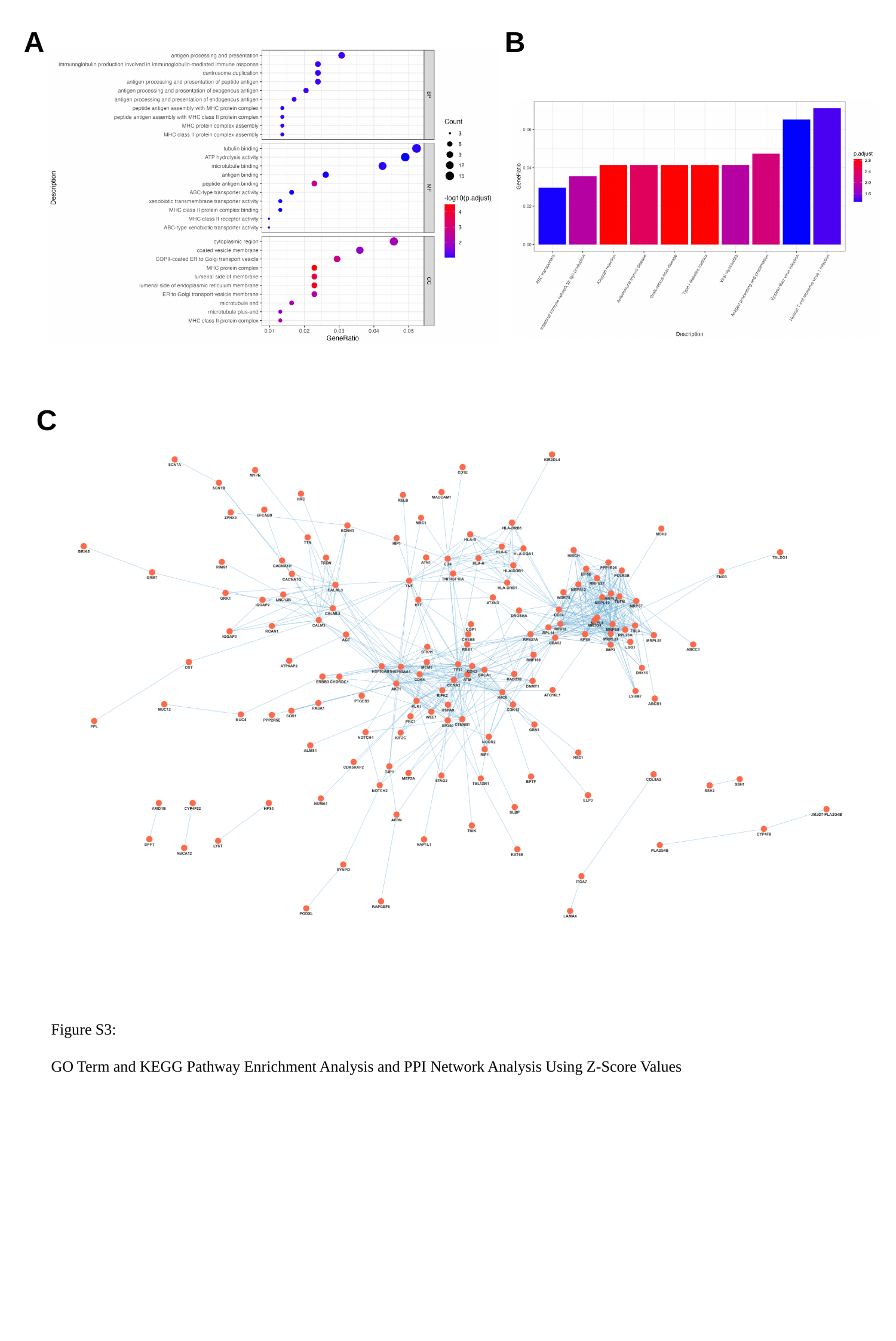

A
B
C
Figure S3:
GO Term and KEGG Pathway Enrichment Analysis and PPI Network Analysis Using Z-Score Values

### Slide 5
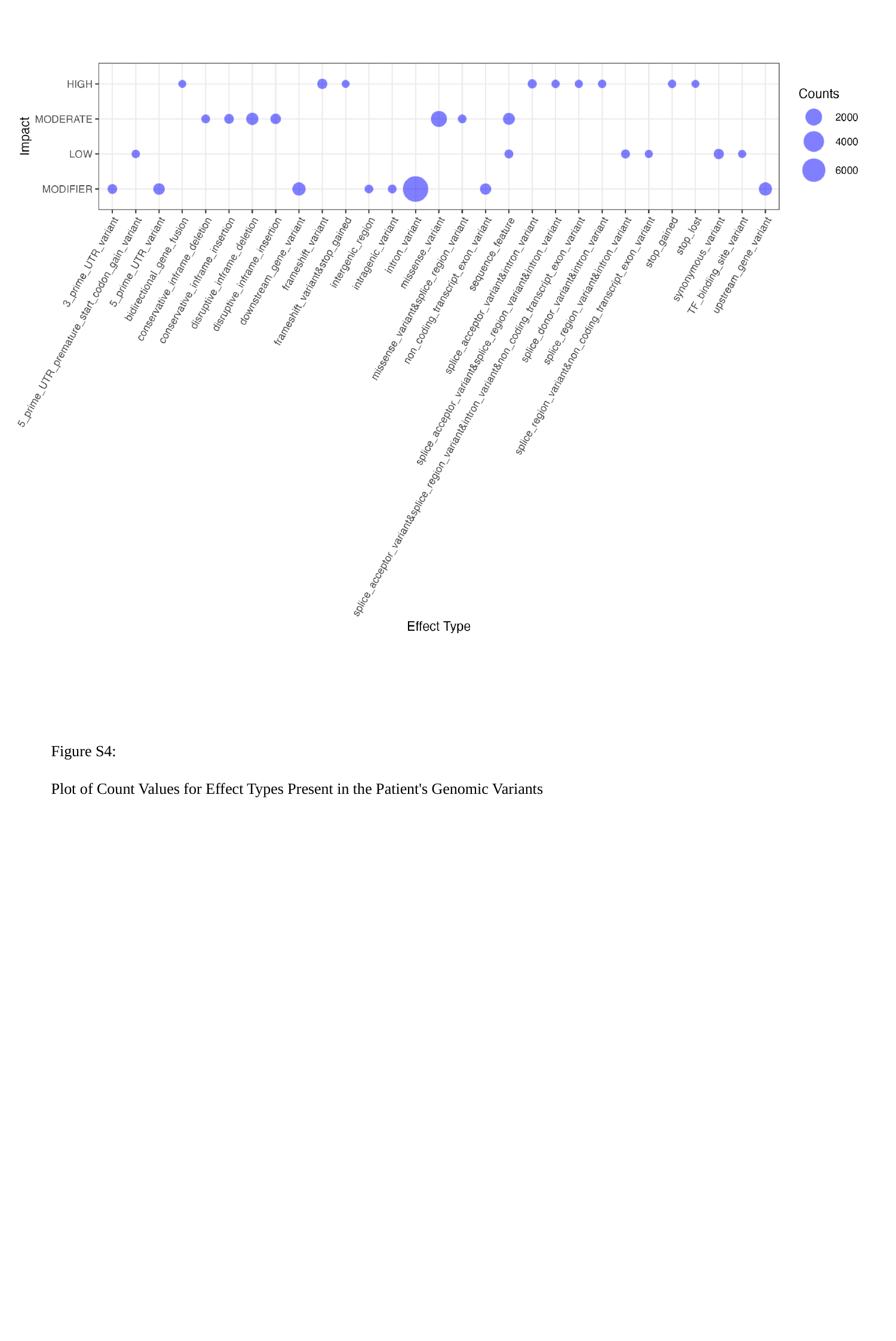

Figure S4:
Plot of Count Values for Effect Types Present in the Patient's Genomic Variants

### Slide 6
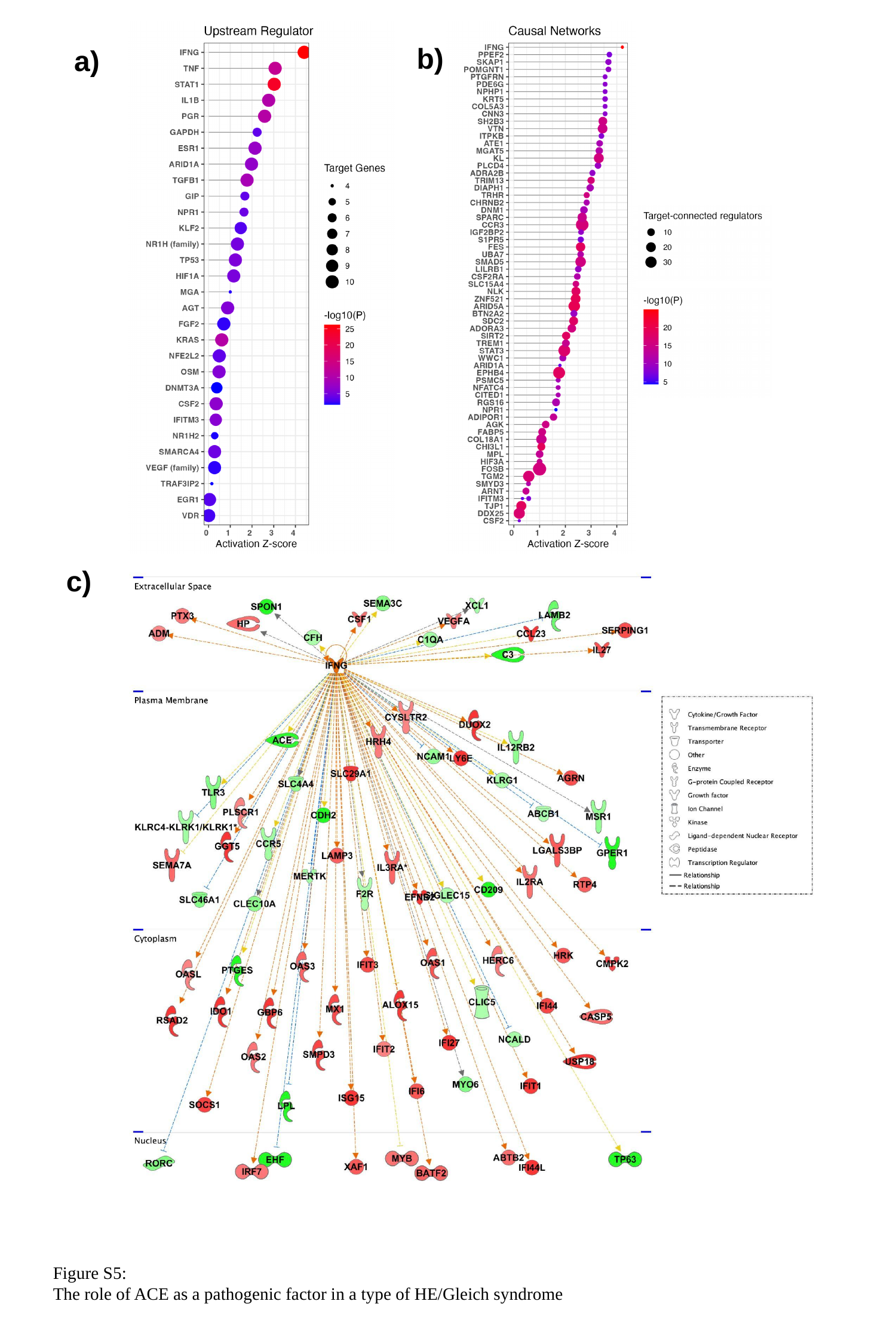

b)
a)
c)
Figure S5:
The role of ACE as a pathogenic factor in a type of HE/Gleich syndrome

### Slide 7
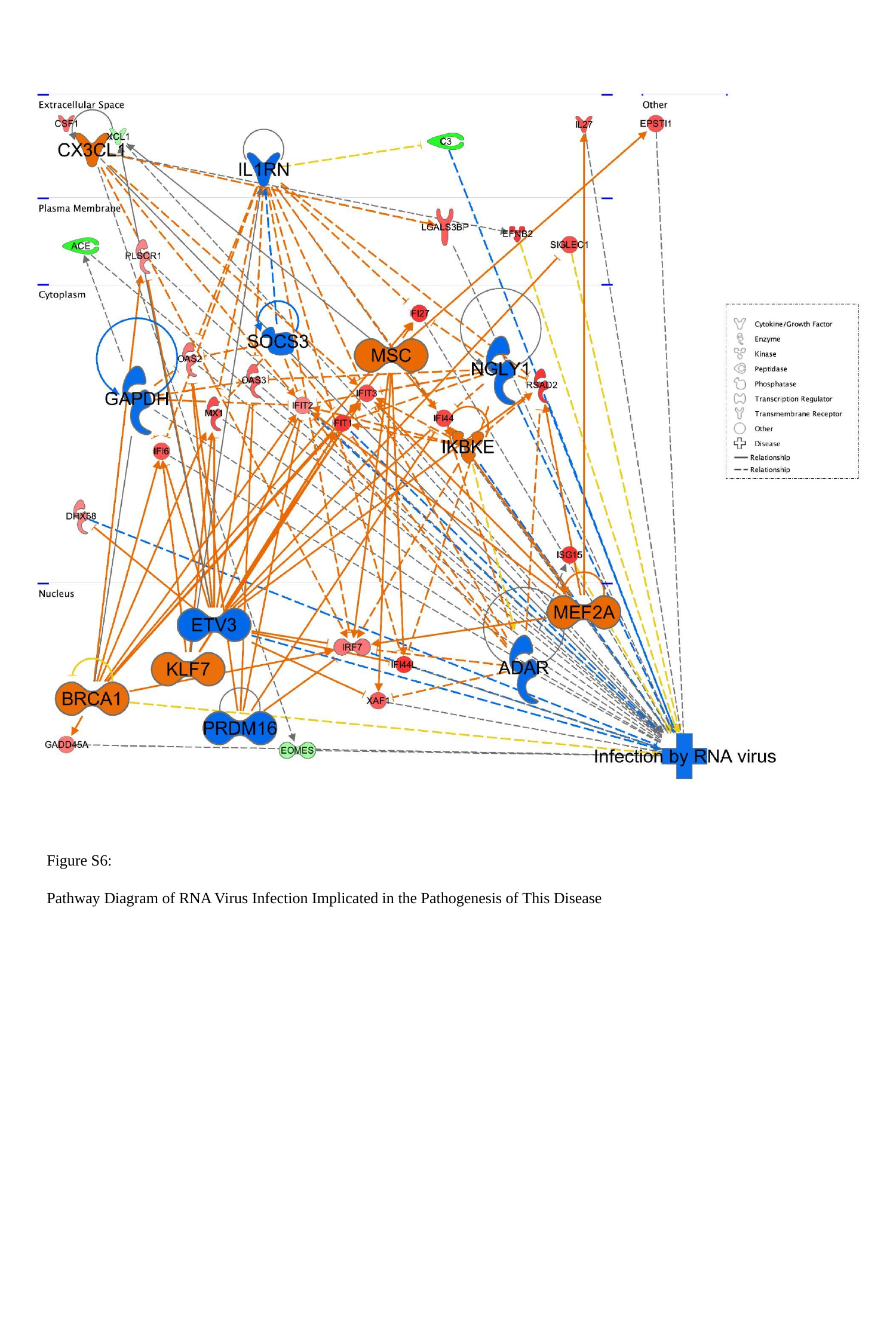

Figure S6:
Pathway Diagram of RNA Virus Infection Implicated in the Pathogenesis of This Disease

### Slide 8
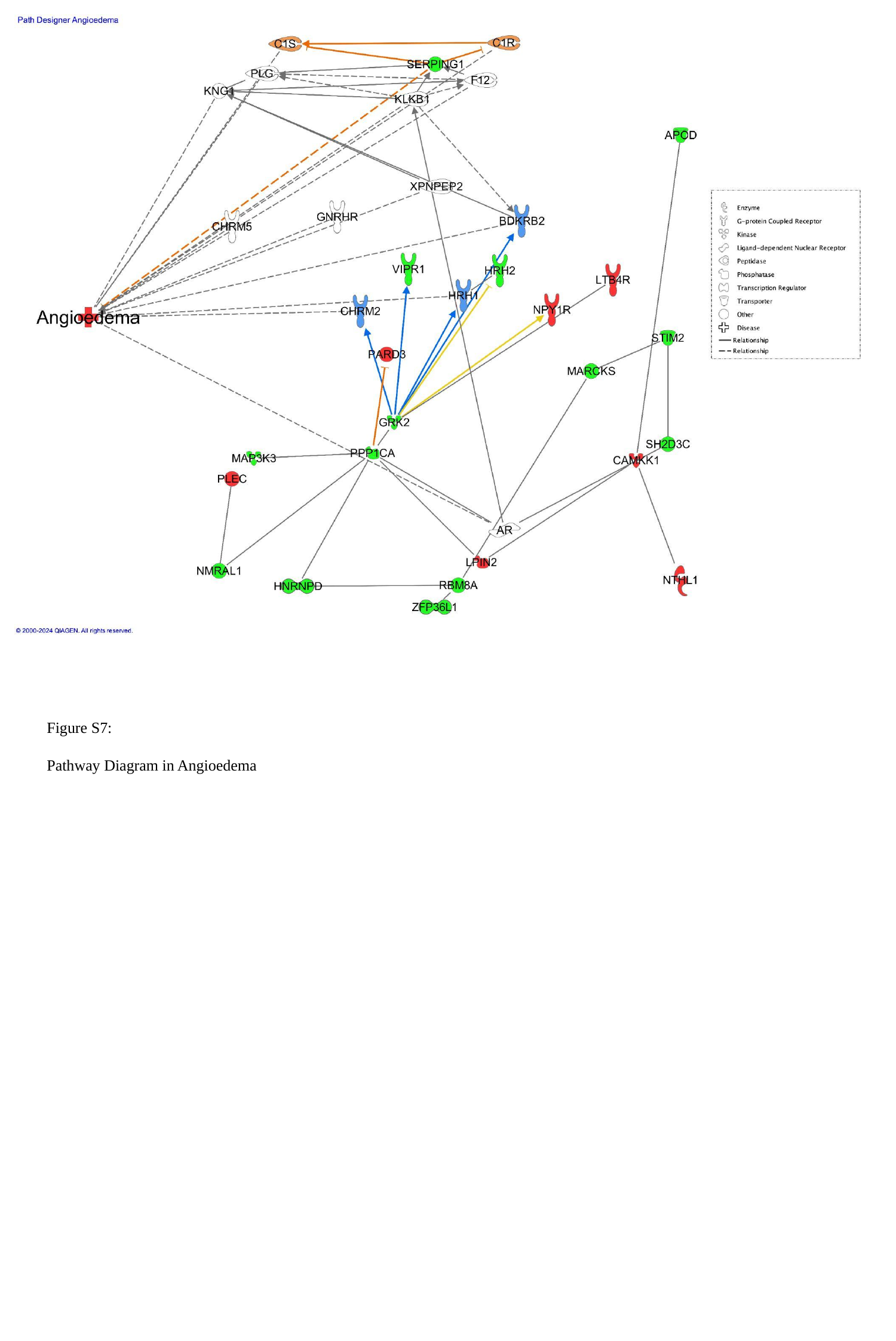

Figure S7:
Pathway Diagram in Angioedema

### Slide 9
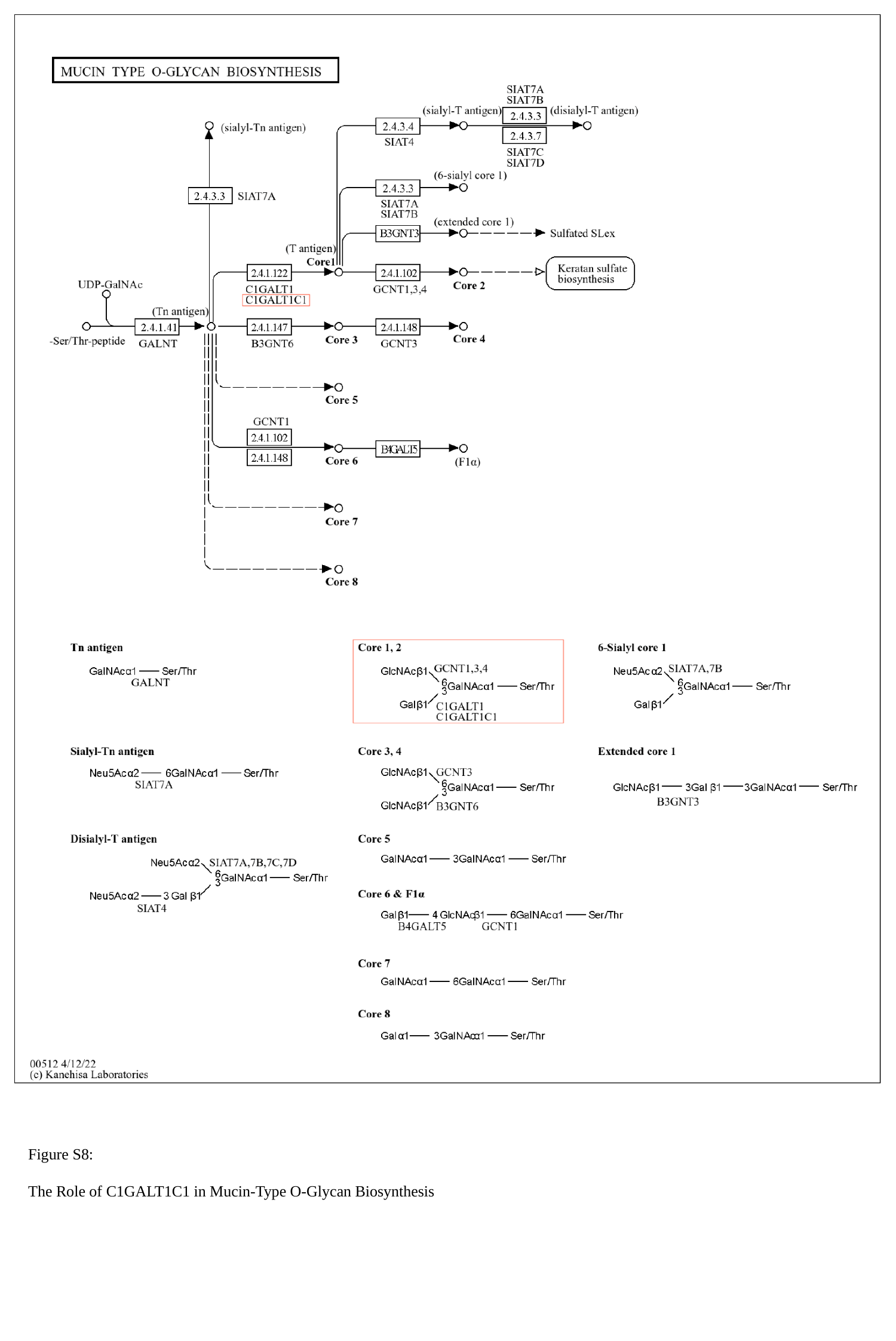

Figure S8:
The Role of C1GALT1C1 in Mucin-Type O-Glycan Biosynthesis
